# The Search for a Diagnostic Indicator of Cochlear Deafferentation: Predicting Age from Auditory Evoked Potential Measures

**DOI:** 10.1101/2025.11.20.25340672

**Authors:** Brad N. Buran, Morgan Thienpont, Sean D. Kampel, Anne E. Heassler, Nicole K. Whittle, Haley A. Szabo, Sarah Verhulst, Naomi F. Bramhall

## Abstract

**Objectives:** Cochlear synaptopathy, a type of cochlear deafferentation that occurs with aging, is expected to be common in humans and to have negative impacts on auditory perception. However, there is currently no means for diagnosing cochlear deafferentation in living humans. Auditory brainstem response (ABR) wave I amplitude and the envelope following response (EFR) are auditory evoked potentials that have been proposed as potential non-invasive indicators of cochlear deafferentation. However, these measures may be impacted by outer hair cell (OHC) dysfunction and changes in endocochlear potential (EP), making them difficult to interpret. One potential method for estimating the degree of deafferentation in individual patients is to combine evoked potential and distortion product otoacoustic emission (DPOAE) measurements with a computational model of the auditory periphery (CMAP). The goal of this study was to evaluate the ability of auditory evoked potentials, with and without the CMAP, to predict age, a risk factor for cochlear deafferentation.

**Design:** In a population with up to a mild sensorineural hearing loss, a CMAP was used with Bayesian regression to predict the frequency-specific degree of deafferentation for individual human participants based on their ABR, EFR, and/or DPOAE measurements and determine whether changes in EP could influence these predictions. Linear regression models were then used to evaluate the ability of the estimated degree of deafferentation and various ABR wave I amplitude, EFR magnitude, and DPOAE measurements to predict age.

**Results:** High frequency ABR wave I amplitude measurements and estimates of deafferentation generated from high frequency ABR wave I amplitudes performed the best at predicting participant age. Accounting for potential changes in OHC function (as indicated by DPOAEs) or the EP in the estimates of deafferentation or by including DPOAEs in the linear regression models had limited impact on the ability of ABR wave I amplitudes to predict age.

**Conclusions:** High frequency ABR wave I amplitudes and estimates of cochlear deafferentation generated from high frequency ABR wave I amplitudes were able to predict participant age within approximately 6 years, with or without incorporating DPOAE measurements. Modeling results indicate that changes in the EP are not a compelling explanation for this outcome. This suggests that high frequency ABR wave I amplitude measurements are good candidates for non-invasive diagnosis of age-related cochlear deafferentation and the CMAP can be used with ABR measurements to predict deafferentation in individual patients.

## Introduction

Both age-related (Schmiedt et al., 1996; Sergeyenko et al., 2013) and noise-induced cochlear synaptopathy (Kujawa & Liberman, 2009), the loss of the synapses between the inner hair cells and the afferent auditory nerve fibers (ANFs), have been described in animal models. Post-mortem temporal bone studies suggest that age-related synaptopathy also occurs in humans (Wu et al., 2019). Despite the limitations associated with post-mortem histology, these studies reflect our best knowledge of synaptic loss in humans due to our inability to image cochlear synapses in living humans, although promising developments may be on the horizon. For example, structural imaging of the vestibulocochlear nerve using structured magnetic resonance imaging (MRI; Harris et al., 2021) shows age-related differences in density, with lower densities associated with older adults. Cochlear synaptopathy is not the only possible explanation for age-related reductions in peripheral auditory function, making it difficult to determine the perceptual consequences of this type of auditory deficit (reviewed in Bramhall & McMillan, 2024).

Outer hair cell (OHC), inner hair cell (IHC), and spiral ganglion cell numbers also decline with age, although temporal bone studies suggest that cochlear synapse loss occurs more rapidly than loss of these other cell types (Wu et al., 2019). Age-related degeneration in the stria vascularis, with an associated decline in the endolymphatic potential (EP), has also been observed in animal models (Gratton et al., 1996; Schulte & Schmiedt, 1992). In humans, temporal bone data demonstrates significant strial degeneration in aged ears (Wu et al., 2020), but an age-related decline in EP has not been confirmed because the EP cannot be directly measured in humans.

Because physiological measures and synapse counts cannot be obtained in the same individuals, there is no consensus on the best physiological measure and stimulus parameters to use for estimating synapse loss. The amplitude of wave 1 of the auditory brainstem response (ABR) and the strength of the envelope following response (EFR) are two non-invasive physiological measures that have been identified in mouse models as possible indicators of cochlear synaptopathy (Kujawa & Liberman, 2009; Parthasarathy & Kujawa, 2018; Shaheen et al., 2015). Although these physiological measures cannot differentiate between cochlear synaptopathy and other types of deafferentation, such as IHC or SGC loss, human temporal bone data suggests that age- and noise exposure-related synaptopathy is the most common type of cochlear deafferentation (Wu et al., 2019; Wu et al., 2021).

ABR wave 1 amplitudes, a direct measure of ANF activity (Hashimoto et al., 1981; Moller & Jannetta, 1981), show progressive age-related declines in mice with histologically confirmed synaptopathy (Sergeyenko et al., 2013) and can be used to predict synapse numbers in mice with age-related or noise-induced synapse loss (Buran et al., 2026). However, there is no consensus on the best ABR stimulus and recording parameters to use for human studies of cochlear deafferentation (reviewed in Bramhall, 2021). Co-occurring outer hair cell (OHC) dysfunction may also complicate interpretation of ABR wave I amplitude as an indicator of deafferentation (Verhulst et al., 2016).

The EFR is a measure of how well the auditory system can encode an amplitude modulated stimulus (Dolphin & Mountain, 1992). Mouse studies suggest that the EFR for a tone sinusoidally modulated at 1000-1024 Hz, believed to target the auditory nerve, is particularly sensitive to synaptopathy (Parthasarathy & Kujawa, 2018; Shaheen et al., 2015). Since the EFR in response to high modulation frequencies is difficult to distinguish from the noise floor in humans (Garrett & Verhulst, 2019; Purcell et al., 2004), most human deafferentation studies use a modulation frequency of 80-120 Hz which results in greater EFR magnitudes (e.g., Bramhall et al., 2021; Mepani et al., 2021; Paul et al., 2017), but is thought to target the auditory brainstem rather than the auditory nerve (Herdman et al., 2002; Kiren et al., 1994; Kuwada et al., 2002). As with the ABR, it is unclear which EFR carrier frequency is the most sensitive to deafferentation. Initial human deafferentation studies used a sinusoidally amplitude modulated (SAM) tone as the EFR stimulus, but a rectangular amplitude modulated (RAM) tone may be more sensitive to deafferentation, result in higher magnitude responses, and be less impacted by OHC dysfunction than a SAM tone due to a sharper stimulus onset (Van Der Biest et al., 2023; Vasilkov et al., 2021). Both SAM and RAM EFR are sensitive to synaptopathy in animal models (Buran et al., 2026; Garrett et al., 2025; Wilson et al., 2021).

A computational model of the human auditory periphery (CMAP; Verhulst et al., 2018) was previously used to predict deafferentation in individual human participants based on their measured ABR wave I amplitudes and distortion product otoacoustic emissions (DPOAEs; Buran et al., 2022).

Predicted deafferentation was correlated with predicted risk factors (noise exposure and age) and perceptual consequences of deafferentation (tinnitus and difficulty with speech perception in noise). The current study expanded on this computational modeling approach by evaluating the ability of both ABR and RAM EFR measures to predict cochlear deafferentation in individual human participants with up to a mild hearing loss. The study also investigated whether changes in EP could explain observed age-related reductions in ABR and EFR measurements. The objective was to determine the optimal physiological measure and stimulus or combination of measures/stimuli for predicting deafferentation in humans.

Given that synapse numbers cannot be confirmed in living humans, age was used as a proxy for deafferentation. Deafferentation predictions generated from different combinations of physiological measures were compared based on their ability to predict participant age.

## Methods

### Participants

Data from 102 adults aged 18-59 years were collected at the National Center for Rehabilitative Auditory Research (NCRAR). Participants were recruited from previous NCRAR studies; fliers posted at the VAPORHCS, Portland area colleges/universities, and in the Portland community. Inclusion criteria included: pure tone air conduction thresholds ≤40 dB HL from 0.25-8 kHz, a normal tympanogram (226 Hz, compliance 0.3-1.5 ml, and peak pressure between ±50 daPa), no air-bone gaps > 15 dB and no more than one air-bone gap of 15 dB, no history of otologic or neurologic disorder (including traumatic brain injury or concussion), and either a native English speaker or learned English prior to age 5. Only individuals meeting all audiological criteria in at least one ear were invited to participate. All participants provided written informed consent and were paid for their participation. All study procedures were approved by the VAPORHCS Institutional Review Board. EFR data from 33 participants were described previously (Bramhall and McMillian, 2024).

### Procedures

Only participants who completed both ABR and EFR testing were included in this analysis. Except for audiometry and tympanometry, test measures were obtained in a single ear. If only one ear met the inclusion criteria, all test measures were collected in that ear. If both ears qualified but there was a factor that would complicate testing in one of the ears, such as cerumen or an ear piercing, the uncomplicated ear was selected as the test ear. In the absence of such a complication, the test ear was determined by rolling a die.

#### Audiometry

Pure tone thresholds were assessed binaurally in all participants from 0.25-8 kHz as part of the screening evaluation. Extended high frequency thresholds from 9-16 kHz were also measured using RadioEar DD450 headphones (New Eagle, PA).

#### Distortion product otoacoustic emissions (DPOAEs)

DPOAE testing was conducted using a custom system consisting of an ER10X probe microphone and EMAV software (Neely & Liu, 1993). DPOAE stimuli were presented at a fixed primary frequency ratio *f_2_/f_1_*=1.2 and responses were obtained using a primary frequency sweep (DPgram) in 1/3 octave increments from 1-8 kHz and 1/6 octave increments from 9-16 kHz at two sets of stimulus frequency levels (*L_1_/ L_2_*): 65/55 dB forward pressure level (FPL) and 50/40 dB FPL. Using in-the-ear calibration, the voltage was adjusted to set *L_1_* and *L_2_* to the desired levels. The ER10X enables FPL calibration, which increases the precision of the stimulus presentation level measurement, particularly for frequencies from 3-7 kHz where interactions of incident and reflected waves produce pressure nodes or standing waves that can cause errors in standard sound pressure level (SPL) calibration (Konrad-Martin et al., 2016). Other than differences in the degree of measurement error, values in FPL and SPL are equivalent. Averaging continued until 48 seconds of artifact-free data were collected, the signal-to-noise ratio (SNR) was ≥ 30 dB, or the noise floor was below -30 dB SPL.

#### Auditory brainstem response (ABR)

ABR data was obtained using an Intelligent Hearing Systems SmartEP system (Miami, FL) and Etymotic Research gold foil ER3-26A tiptrode electrodes (Elk Grove Village, IL) placed in the ear canal. Alternating polarity toneburst stimuli (1, 2, 2.8, 4, 5.6, or 8 kHz) were presented at 100 and 110 dB peak-to-peak equivalent SPL (peSPL) with the exception of 8 kHz, which was presented at 100 and 105 dB peSPL due to the output limitations of the transducer. Toneburst durations were chosen to maximize frequency specificity and brevity: 4 ms for 1 kHz, 3 ms for 2 kHz, 2.5 ms for 2.8 kHz, 2 ms for 4 kHz, 1.43 ms for 5.6 kHz, and 1 ms for 8 kHz. For the 100 dB peSPL stimuli, 2048 sweeps were averaged, while only 1024 sweeps were averaged for the 105 dB peSPL and 110 dB peSPL stimuli to limit exposure to high intensity levels. The stimulus repetition rate was 11.1/sec and at least two waveform replicates were obtained. Electrode impedance was ≤ 5.0 kΩ for all participants. Wave I peaks and the following negative troughs were scored by two experienced audiologists using a peak picking program (adapted from Buran, 2015). Disagreements were resolved by a third audiologist. Wave I amplitude was defined as the voltage difference between the positive peak and the following negative trough (Supplemental Figure 1A).

#### Envelope following response (EFR)

EFRs were recorded in a sound booth using Intelligent Hearing Systems SmartEP-CAM (Miami, FL) and electrostatically shielded Etymotic Research ER-3A insert transducers (Grand Prairie, TX). Participants were seated in a comfortable recliner in the reclined position, asked to close their eyes, and encouraged to sleep. The active electrode was placed at the vertex (Cz), the ground on the low forehead (Fpz), and the reference on the mastoid of the test ear. Electrode impedance was ≤ 5.0 kΩ for all but three participants, where impedance was ≤ 9.3 kΩ. Stimuli consisted of 70 dB peSPL RAM tones 100% modulated at 110 Hz with a 25% duty cycle and a 2.5% Tukey window applied to the onset and offset of each individual cycle of the RAM tone (Vasilkov et al., 2021). Carrier frequencies were 1, 2, and 4 kHz. Stimuli were 500 ms in duration, presented in alternating polarity, and each recording consisted of 1002 sweeps. To minimize entrainment of evoked potentials to the stimulus, the interstimulus interval was uniformly jittered between 100 and 120 msec. EFR magnitude was calculated similarly to the bootstrapping approach described in Zhu et al. (2013). Four hundred trials were drawn with replacement (200 from each polarity), averaged, and the power spectrum was computed (Supplemental Figure 1B). This process was repeated 100 times to generate a distribution of the power for each frequency bin. The average value of the distribution at the modulation frequency and the first four harmonics was used to estimate the raw EFR response. To estimate the noise floor, the power in the fourth to seventh discrete Fourier transform (DFT) bins on either side of the frequency of interest was averaged.

#### Computational modeling of cochlear synapses

A combination of DPOAE, ABR, and EFR measurements was used to predict the number of functional auditory nerve fibers per IHC in individual human participants across cochlear frequency (i.e., the synaptogram). Aside from loss of cochlear synapses, the synaptogram also captures other forms of damage (e.g., IHC and SGC loss) that result in reduced input from the auditory periphery; thus, the synaptogram can be thought of as a measure of cochlear deafferentation. A previous approach, described in Buran et al. (2022), used DPOAE and ABR data to predict each participant’s synaptogram. In this report, that approach was expanded to also incorporate EFR measurements. Throughout this report, **bold** symbols represent vectors. Thus, ***θ*** represents a vector and *θ*_[0]_ represents an element of that vector.

Although ABR wave I amplitude and EFR magnitude are used as indicators of cochlear synaptopathy, evoked responses can also be affected by decreased cochlear gain (e.g., due to OHC dysfunction). For this reason, the computational approach must account for potential OHC dysfunction when generating predictions of synaptic loss. The CMAP includes stages for cochlear mechanics and spiking activity of ANFs and brainstem nuclei. By altering the parameters of the cochlear mechanics stage, the CMAP can simulate spiking responses in an ear with OHC dysfunction. Wave I of the ABR can be generated by multiplying the simulated ANF responses by the predicted number of synapses per IHC at each characteristic frequency (CF; i.e., synaptogram) and then summing the ANF responses. Since the brainstem is thought to contribute to the EFR at low modulation frequencies, simulated ANF responses are passed through models for the cochlear nucleus (CN) and inferior colliculus (IC). The ANF, CN, and IC responses are then summed to generate the EFR.

The CMAP generates simulations of low, medium and high spontaneous rate (SR) ANFs, enabling incorporation of these fiber subtypes into the ABR simulations. Although Buran et al. (2022) assumed that first all low, then all medium, and finally high SR ANFs are lost based on work from animal studies (Furman et al., 2013; Schmiedt et al., 1996), this assumption may not hold true. Computational modeling studies of the EFR indicate that a subset of high SR fibers is likely lost before all low and medium fibers (Encina-Llamas et al., 2019), and single-unit recordings in mice did not find evidence for selective loss of low SR fibers (Suthakar & Liberman, 2021). Accordingly, in this analysis, remaining ANFs were distributed across SR according to a fixed ratio based on data from cats (16% low, 23% medium, and 61% high; Liberman, 1978).

When fitting the CMAP to the DPOAE, ABR, and EFR data, two parameters were adjusted to minimize the difference between simulated and measured data: the pole function (***α***) and the synaptogram (***θ***). The pole function represents the active cochlear amplification process, impacting the overall gain that is fed into the IHC stage of the model. The synaptogram represents the average number of functional auditory nerve fibers per IHC as a function of frequency, which affects the overall amplitude of auditory evoked response. Only the middle ear filtering and cochlear mechanics stages of the CMAP are required to predict DPOAEs. Thus, the model fitting was split into multiple steps. The first step predicted the participant-specific pole function required by the cochlear mechanics stage based on measured DPgrams (Keshishzadeh & Verhulst, 2021). The participant-specific pole function was then frozen and the CMAP was used to simulate participant-specific ANF responses for the ABR and EFR stimuli used in this study. Bayesian regression was then used to estimate the synaptogram.

Separate models were fit using various subsets of the data (e.g., only 4 kHz ABR from all participants, only 4 kHz EFR, both 4 kHz ABR and 4 kHz EFR, etc.). All models were fit using Bayesian regression to maximize the Normal likelihood of free parameters using pymc (Salvatier et al., 2016). The prior for the standard deviation was set to Half Cauchy with a scale parameter (beta) of 1. Each model was fit four times for 2000 samples following a 1000 sample burn-in period using the No U-Turn Sampler (Hoffman & Gelman, 2014). Posterior samples were combined across all fits for inference. Gelman-Rubin statistics were computed to ensure that the four fits, each of which started with a random estimate for each parameter, converged to the same final estimate (*̂R* < 1.1).

##### 1. Correcting for DPOAEs

The CMAP simulates basilar membrane (BM) vibration using a series of longitudinally coupled cochlear mechanical filters, each centered at a given cochlear frequency. The effects of OHC dysfunction (i.e., broader cochlear filtering and reduced sensitivity) can be simulated by reducing the gains of these filters (i.e., the pole function). Thus, the pole function, ***α***, can be adjusted in a CF-dependent manner to reflect frequency-specific OHC damage in individual human participants (Verhulst et al., 2012; Verhulst et al., 2016). First, a dataset of 20,000 synthetic pole functions, corresponding to OHC dysfunction profiles from fully intact to severely damaged, were used to simulate DPOAE data for the DPOAE frequencies used in this dataset. Unlike our previous proof-of-concept implementation for predicting the participant-specific pole function (Buran et al., 2022), which used DPgrams generated for a single level (*L_2_* = 55 dB SPL) for standard audiometric frequencies (1-8 kHz), we expanded our approach to include two DPOAE levels (*L_2_* = 40 and 55 dB SPL) across standard and extended high frequencies (9-16 kHz), added imputation for missing data, greatly expanded the training space (from 26 simulated profiles to 20,000 profiles), and implemented a deeper neural network with dropout regularization (forcing the network to find redundant pathways to arrive at the same conclusion, ensuring that no single neuron becomes too dominant) and early stopping (which prevents overtraining).

To ensure that the training space captured both realistic clinical phenotypes and broad physiological variability, the pole functions were constructed using two distinct computational strategies. The first 10,000 samples were generated using a Dirichlet mixing approach to simulate clinically plausible hearing loss profiles. For each synthetic participant, between one and three base clinical profiles were randomly selected from a predefined set and blended using sparse convex combinations with mixing weights drawn from a Dirichlet distribution to ensure the coefficients summed to one. The second 10,000 samples were generated using a randomized mathematical approach which introduced broader physiological variations into the dataset, ensuring that the neural network would not overfit to specific clinical archetypes and could generalize to anomalous or highly irregular cochlear gain patterns.

A deep neural network was then trained on the simulated DPOAE dataset. The network architecture consisted of a 32-neuron input layer, four hidden layers comprising 256, 512, 512, and 256 neurons with interspersed dropout regularization, and a 1001-neuron output layer. This trained network was then used to predict participant-specific pole functions, ***α_i_***.

##### 2. Priors for Synapse Counts

As in Buran et al. (2022), the predicted synapse count per IHC for participant *i* at position *g* was modeled as *N*_i_(*g*) = exp (−exp(*θ*_i[0]_)(*g* − *θ*_i[1]_)^2^ + *θ*_i[2]_). Participant *i*’s maximum number of synapses per IHC was exp(*θ*_i[2]_), the vertex was positioned at cochlear frequency *g* = *G*(*θ*_i[1]_), and the curvature of the parabola at the vertex was defined by 2 ⋅ exp(*θ*_i[0]_). *G*(*g*) is the Greenwood equation (Greenwood, 1990), converting *g* to the cochlear frequency. The vector of participant-specific parabola coefficients, ***θ_i_***, were assumed to be normally distributed around a population mean vector, Θ, with covariance matrix, **Σ**. Synapse counts per IHC from a set of 19 human cadaver ears (Wu et al., 2019) were used to generate priors for Θ and **Σ**, ensuring that the model provided realistic predictions of the synaptogram.

##### 3. Predicting Deafferentation

As in Buran et al. (2022), the CMAP was used to predict the instantaneous firing rate for high, medium and low SR ANFs distributed across the tonotopic axis of the human cochlea. The ABR response was calculated from the summed response of the simulated ANF activity scaled by the predicted number of synapses at that cochlear frequency as described by Equation 3 in Buran et al. (2022) with one change. In Buran et al. (2022), simulated ABRs had a sex-specific scaling factor, exp(*c*_0_ + *c*_1_ ⋅ *S*_i_), where *c*_0_ corrects for differences between simulated and measured ABR wave I amplitude, *c*_1_ is the difference between males and females, and *S*_i_ = 0 if the participant is male, 1 if the participant is female. Because simulated ABRs did not recapitulate measured frequency-specific differences in the scale of ABR wave I amplitude (Supplemental Figure 1A), *c*_0_ in Equation 3 of Buran et al. (2022) was replaced by a frequency-specific scaling constant, *c*_0,ƒ_, to correct for differences between the simulated and measured waveform at frequency *f*, providing a frequency and sex-specific scaling factor for the ABR, exp(*c*_0,ƒ_ + *c*_1_ ⋅ *S*_i_).

This scaling constant was assigned a Normal prior with the mean set to the natural logarithm of the ratio of measured to simulated ABR wave I amplitudes for the corresponding frequency and the standard deviation was set to 1.

Similar to the ABR response, the EFR response, EFR_i_(*f*), for participant *i* was modeled as the sum of the unitary contribution of each fiber to the overall EFR magnitude:

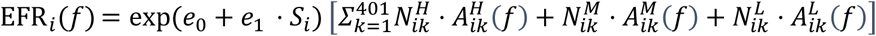

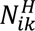 is the average number of high SR fibers at the characteristic frequency indexed by *k* for participant *i*. 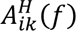 (*f*) is the unitary contribution to the EFR of a high (*H*) SR ANF for participant *i* with characteristic frequency indexed by *k* responding to an EFR stimulus of frequency *f*. Medium (*M*) and low (*L*) SR fibers are similarly represented. The unitary contribution is represented as a complex number, ensuring that phase information is preserved. Unlike the ABR, the ratio between simulated and measured EFRs was constant across frequency (Supplemental Figure 1B), so a frequency-specific scaling factor was not used. Thus, *e*_0_ represents the scaling constant needed to correct for differences between the simulated and measured magnitude. The mean of the prior was set to the logarithm of the average ratio of the measured to simulated EFR magnitude across carrier frequency and the standard deviation was set to 1. Since there may be sex differences in EFR magnitude (Krizman et al., 2012), a sex-specific scaling constant, exp(*e*_0_ + *e*_1_ ⋅ *S*_i_), was included. For the difference between females and males, *e*_1_, a Normal prior with mean of 0 and standard deviation of 1 was used. All other parameters were represented similarly to the ABR response.

##### 4. Predicting endolymphatic potential

The CMAP incorporates a biophysical IHC-auditory nerve stage which models the effect of the EP on the current through the mechanoelectric transduction channel (Equation A4 in Verhulst et al. 2018). Simulated ABR wave I amplitudes and EFR magnitudes were generated from the CMAP while varying the EP between -40 and 120 mV. The relationship between simulated ABR wave I amplitude and EP was fit to the equation R_ABR,ƒ,l_(EP) = *y*_floor_ + *A*[ln(1 + *e*^k(EP–EP0)^)]^y^ where R (EP) is the change in ABR wave I amplitude relative to the simulated amplitude for EP=90 mV where f is the ABR stimulus frequency and l is the ABR stimulus level. Stimulus and participant-specific ABRs were scaled by this ratio ABR_i_(*f*) ⋅ *R*_ABR,ƒ,l_(EP_i_). The relationship between simulated EFR magnitude and EP was represented similarly, and denoted as R_EFR,ƒ_(EP). When generating predictions of ABR and/or EFR, the Bayesian regression model incorporated a participant-specific coefficient for EP, *EP*_i_, which was assigned a relatively uninformative Normal prior with mean 55 and standard deviation 15 given the inability to directly measure EP in living humans.

### Statistical Analysis

#### Reanalysis of published mouse data

To validate the approach presented in this report of using the ability to predict age as a proxy for the ability to predict deafferentation, mouse data from Buran et al. (2025) was re-analyzed to determine how well synapse counts perform at predicting a mouse’s age (in weeks). For this analysis, noise-exposed mice were excluded. The distributions of measured DPOAE thresholds, ABR wave 1 amplitudes, and synapse counts are shown in the young and aged groups of Figure 1 of Buran et al. (2025). To test the relative ability of synapse counts to predict age, a linear regression model was constructed, *y*_i_ = *β*_0_ + *β*_1_ ⋅ ln x_i[ƒ]_ + *ε*_i_, where *y*_i_ is the age of the *i*-th mouse and the mouse-specific synapse count at frequency *f* is represented by x_i[ƒ]_. Model performance was assessed using twenty repeats of 20-fold cross-validation as described in *Assessing model performance* in the *Methods*.

**Figure 1.**
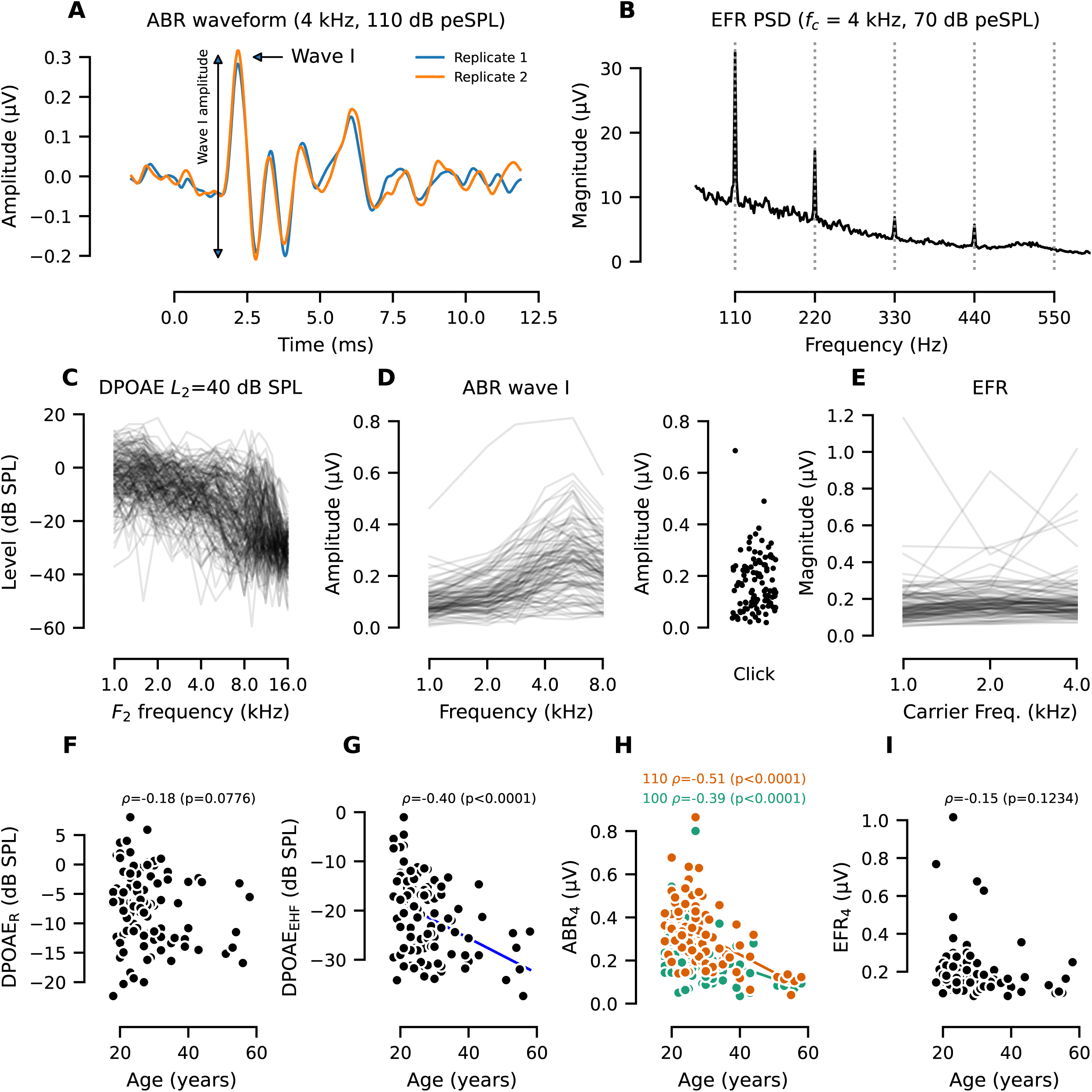
The sample generated a large range of DPOAE, ABR, and EFR measurements which were correlated with age. Exemplar ABR waveform for a 4 kHz, 110 dB peSPL toneburst illustrating how ABR wave I amplitude was measured (**A**). Exemplar magnitude spectrum for a 4 kHz RAM EFR showing the fundamental (110 Hz) and four harmonics that were summed to calculate the EFR magnitude for each participant (**B**). (**C-E**) Plots show variation across individual participants (N=102) in DPOAE levels (**C**), ABR wave I amplitudes (**D**), and RAM EFR magnitudes (**E**). DPOAE levels for *f_2_* < 9 kHz (DPOAE_R_; **F**) were not correlated with age. Average DPOAE levels for *f_2_* ≥ 9 kHz (DPOAE_EHF_; **G**) and ABR wave I amplitudes for a 4 kHz toneburst (ABR_4_) for both 100 (green) and 110 (orange) peSPL stimuli (**H**) were correlated with age. RAM EFR magnitudes for a 4 kHz carrier (EFR_4_) were not correlated with age (**I**). For all DPOAEs, *L_2_* = 40 dB SPL (**C, F, G**). Individual lines or markers indicate data from individual participants. For **F-I**, Pearson’s correlation coefficients and p-values are shown above each plot. A solid regression line is shown only for p < 0.05.

To more closely parallel the linear regression models used to predict age from the human synaptogram coefficients, mouse synaptograms were fit to Equation 1 to obtain mouse-specific synaptograms (***θ_i_***) and then age was predicted using the linear regression model *y*_i_ = *β*_0_ + 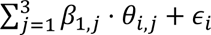

#### Models for predicting participant’s age

To test the relative ability of the synaptogram to predict the age of each participant, linear regression models were constructed based on participant *i*’s synaptogram, ***θ_i_***, as *y*_i_ = *β*_0_ + 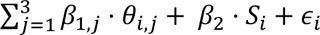. The age of participant *i* is represented by *y*_i_, *θ*_i,j_ is the *j*-th parameter of the participant’s synaptogram, and the participant-specific sex is represented as *S*_i_, where *S*_i_ = 0 if the participant is male, 1 if the participant is female. The residual error term is represented by *ε*_i_. It was not necessary to include DPOAEs as predictors in the model since the CMAP corrects for DPOAEs. Although synaptograms were corrected for sex, sex was included as a predictor in all models to facilitate comparisons with ABR, EFR, and DPOAE measurements because there are reported sex differences for ABR and EFR (Krizman et al., 2012; Trune et al., 1988), and the average age of the female participants in the sample differed from the average age of the male participants by 10 years.

A linear regression model, 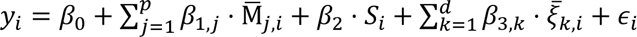 was used to test the ability of each raw evoked potential measurement (ABR, EFR, or DPOAE) to predict participant age. The age of participant *i* is represented by *y*_i_, the participant-specific evoked potential measure *j* is represented by Μ_j,i_, the number of evoked potentials is represented by *p*, the participant-specific DPOAE measure *k* is represented by ξ_k,i_, the number of DPOAE measurements is represented by *d*, and the residual error term is represented by *ε*_i_. Since the raw ABR data had two stimulus levels per frequency, each stimulus level was treated as a separate predictor in the model and an interaction term was also included (e.g., *p* = 3 when using only ABR wave I amplitude at 4 kHz to predict age).

To establish a baseline for interpreting the prediction error, an intercept-only model (i.e., *y*_i_ = *β*_0_ + *β*_2_ ⋅ *S*_i_ + *ε*_i_) was tested in which the predicted age was the average age of all participants of the same sex. In addition, because extended high frequency DPOAEs are correlated with age (e.g., Hauser et al., 2025; Mepani et al., 2021), the ability of the DPOAE measurements to predict age was also evaluated (i.e., 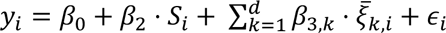). An ideal metric for assessing deafferentation would perform better at predicting age than the intercept-only and the DPOAE-only models. Since extended high frequency audiometric thresholds (9-16 kHz) are strongly correlated with age (Hauser et al., 2025), the ability of these measurements to predict age was also evaluated to provide a reference for discussion.

#### Assessing model performance

Model performance was assessed with 20-fold cross-validation using statsmodels (Seabold & Perktold, 2010). To ensure robust estimates, the 20-fold process was repeated 20 times with random shuffling (Burman, 1989). Performance was quantified using root-mean-squared error (RMSE) for age.

#### Computational simulations of ABR wave I amplitude and EFR magnitude

To test the relative impact of OHC dysfunction and cochlear deafferentation on the ABR and EFR, the CMAP was used to simulate ABR and EFR responses under different hearing loss conditions. To simulate the impact of high frequency OHC dysfunction, the exemplar pole functions provided with the Verhulst et al. (2018) model associated with various high frequency hearing loss slopes (ranging from 5 to 35 dB) were used. To generate realistic deafferentation profiles as a function of age, the Wu et al. (2019) dataset was fit using Equation 2 in Buran et al. (2022), which allows the coefficients of the synaptogram to vary as a linear function of age. The fitted equation was then used to generate the expected synaptogram for a human at each decade from 0 (neonate) to 60 years of age under the assumption that low, medium, and high SR fibers have an equal chance of being lost. These pole functions and synaptograms were then used as inputs for the CMAP to generate simulated ABR wave I amplitudes and EFR magnitudes.

#### Assessing the impact of endolymphatic potential

To assess the ability of models with and without EP to predict participant age, the CMAP was fit to combined ABR wave I amplitude at 4 kHz and RAM EFR magnitude at 4 kHz. Four variations of this model were tested: 1) a baseline-only model in which both EP and deafferentation were frozen, 2) an EP-only model in which deafferentation was frozen, 3) a deafferentation-only model in which EP was frozen, and 4) a model in which both EP and deafferentation were fitted parameters. To determine the ability of each model to predict participant age, the following models were used: *y*_i_ = *β*_0_ + *β*_3_ ⋅ *S*_i_ + *ε*_i_ for the baseline model, *y*_i_ = *β*_0_ + *β*_1_ ⋅ *EP*_i_ + *β*_3_ ⋅ *S*_i_ + *ε*_i_ for the EP-only model, *y*_i_ = *β*_0_ +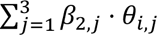 + *β*_3_ ⋅ *S*_i_ + *ε*_i_ for the deafferentation-only model, and 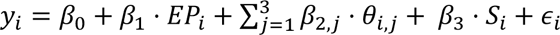 for the EP and deafferentation model.

## Results

### Measurement characteristics and relationships with age

The DPOAE, ABR (Figure 1A), and EFR data (Figure 1B) collected from 102 participants (30 males and 72 females; 18 to 58 years) represents a wide range of cochlear gain loss (OHC dysfunction as indicated by DPOAEs; Figure 1C), ABR wave I amplitudes (Figure 1D) and EFR magnitudes (Figure 1E). To investigate relationships between DPOAEs and ABR/EFR, the DPOAE data were split by frequency into regular (DPOAE_R_, f_2_ < 9 kHz) and extended high frequency (DPOAE_EHF_, f_2_ ≥ 9 kHz) DPOAEs and the DPOAE levels for *L_2_* = 40 dB SPL were averaged across frequency. DPOAE_EHF_ and ABR wave I amplitude for a 4 kHz toneburst (ABR_4_) were correlated with age, but DPOAE_R_ and EFR magnitude for a 4 kHz carrier (EFR_4_) were not (Figure 1F-I). Although the majority of participants had normal hearing thresholds (≤ 20 dB HL from 0.25-8 kHz), seven participants had a mild hearing loss (at least one audiometric threshold of 25-40 dB HL from 0.25-8 kHz).

### Simulated versus measured DPgrams, ABR wave I amplitudes, and EFR magnitudes

The ABR, EFR, and DPOAE measurements were used to predict the degree of cochlear deafferentation in each participant by fitting the CMAP to various combinations of the evoked potential measures. The first step of fitting the CMAP was to individualize the cochlear gain stage (specifically, the pole function which is an indicator of OHC function), for each participant. Using a neural network model trained on simulated DPgrams, a pole function was predicted for each participant and a simulated DPgram was generated from the predicted pole function (Figure 2A-C). Simulated and measured DPOAE levels were well matched (e.g., *f_2_* = 4 kHz, Figure 2D) for both low (*L_2_* = 40 dB SPL) and high (*L_2_* = 55 dB SPL) intensity levels, with strong correlations (Figure 2E), and average prediction errors of 7.5 dB SPL or less across all *f_2_* frequencies (Figure 2F).

**Figure 2.**
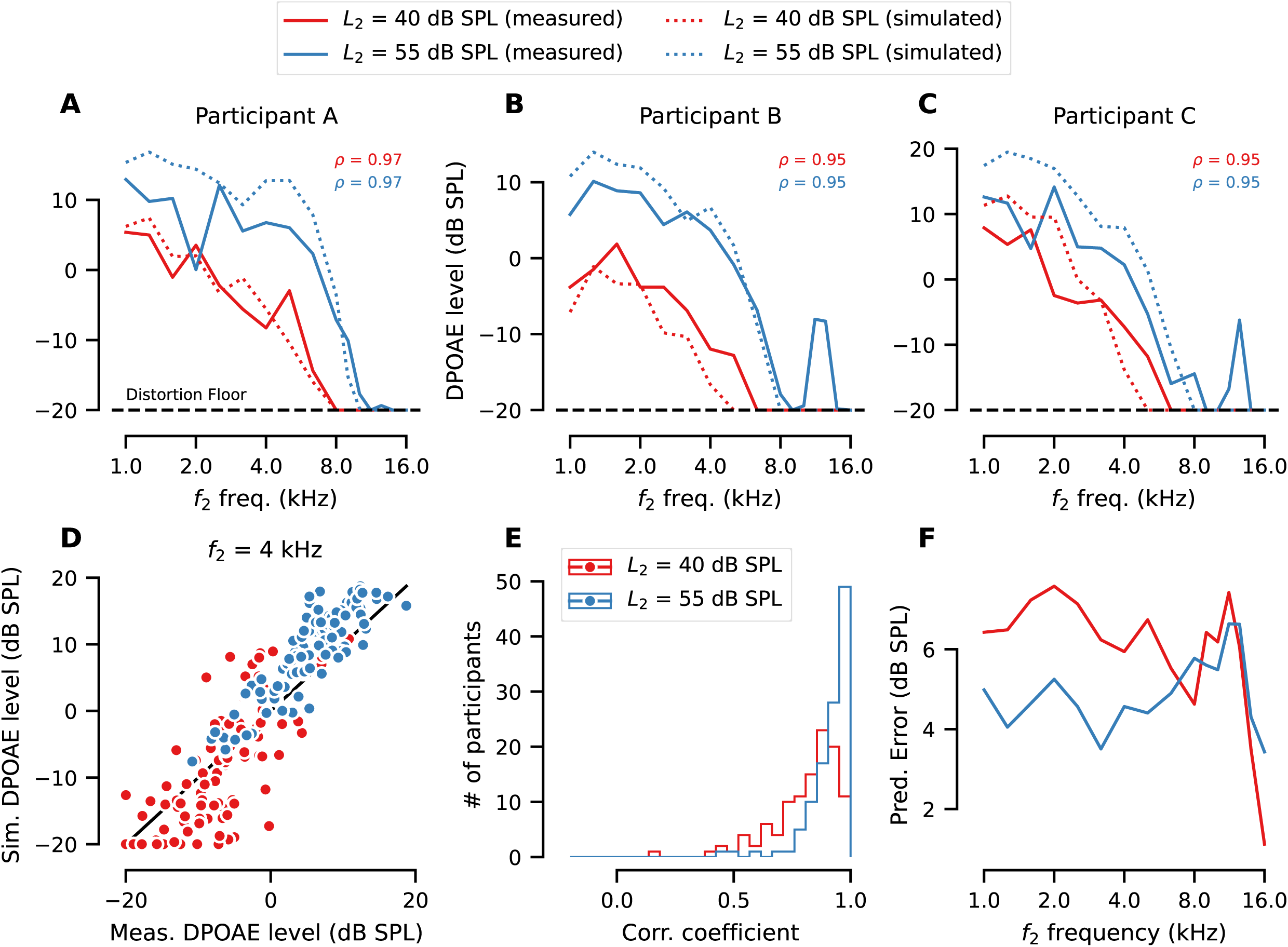
Simulated DPgrams match observed DPgrams. (**A-C**) Each panel shows the relationship between the simulated DPgram (dotted line) and the measured DPgram (solid line) for an exemplar participant. DPgrams for low (*L_2_* = 40 dB SPL, red) and high (*L_2_* = 55 dB SPL, blue) *L_2_* levels are shown separately. Black dashed line indicates the distortion floor of the DPOAE measurement system. The correlation coefficient between each simulated and measured DPgram is shown for both low-(red) and high-level (blue) DPgrams. (**D**) For an *f_2_* of 4 kHz, simulated DPOAE levels match measured DPOAE levels for both a low- and high-level stimulus. Points indicate data from individual participants. Solid black line indicates unity. (**E**) Histograms of the correlations between simulated and measured DPgrams, broken down by low- vs. high-level stimuli. (**F**) Prediction error was calculated as the root mean squared difference between the simulated and measured DPOAE levels. Average prediction error across all participants is shown separately for each *f_2_* and *L_2_*. The drop in prediction error for *f_2_* = 16 kHz is due, in part, to the large number of censored observations at this frequency (i.e., measurements which were below the distortion floor of the system).

Once the CMAP was individualized for each participant, Bayesian regression was used to fit the CMAP to various combinations of each participant’s measured ABR wave I amplitudes and EFR magnitudes. Based on prior histological work in human temporal bones (Wu et al., 2019), the synaptogram was modeled as a parabola and the fits were constrained by the known range of human synapse counts. Synaptograms were predicted using only ABR_4_ (Figure 3A), only EFR_4_ (Figure 3B), or both ABR_4_ and EFR_4_ (Figure 3C). To validate how well the predicted synapse counts simulated the evoked potentials, the CMAP (individualized using the participant-specific pole function and synaptogram) was used to simulate ABR wave I amplitude and EFR magnitude. The simulations were compared to the measured ABR wave I amplitude (Figure 3D-F) and EFR magnitude (Figure 3G-I). When fit to only ABR_4,_ simulated ABR wave I amplitudes were highly correlated with measured ABR wave I amplitudes (Figure 3D); however, simulated EFR magnitudes were not correlated with measured EFR magnitudes (Figure 3G). When fit to only EFR_4_, simulated EFR magnitudes were correlated with measured EFR magnitudes (Figure 3H); however simulated ABR wave I amplitudes were not correlated with measured ABR wave I amplitudes (Figure 3E). When fit to both ABR_4_ and EFR_4_, simulated and measured ABR wave I amplitudes were correlated (Figure 3F), but not simulated and measured EFR magnitudes (Figure 3I).

**Figure 3.**
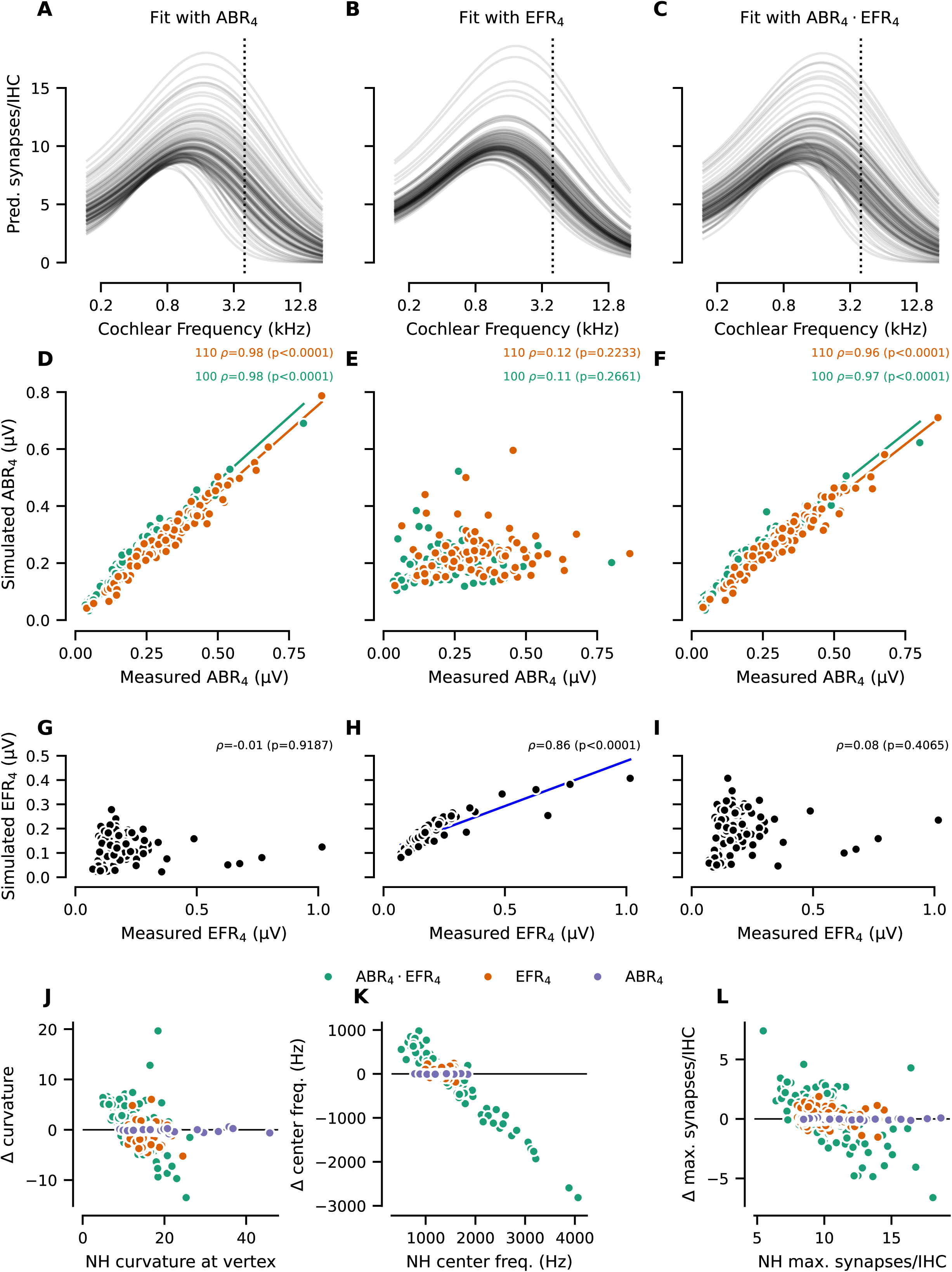
Predicted synapse counts and simulated evoked potentials after individualizing the CMAP using the DPgram. Predicted synaptograms (number of synapses per IHC as a function of cochlear frequency) generated using ABR wave I amplitude at 4 kHz (ABR_4_, **A**), RAM EFR magnitude for a 4 kHz carrier (EFR_4_, **B**), or both 4 kHz ABR and EFR (ABR_4_ · EFR_4_, **C**). For **A-C**, lines represent individual participants and vertical dashed line indicates the stimulus frequency. Simulated vs. measured ABR wave I amplitudes (**D-F**) and EFR magnitudes (**G-I**) are plotted for a 4 kHz stimulus based on the individualized synaptogram of each participant as shown in **A-C**. For **D-I**, dots indicate individual participants. Pearson’s correlation coefficients and p-values are shown above each plot. A solid regression line is shown only for p < 0.05. To assess the impact of cochlear gain loss on predicted synaptograms, the synaptogram coefficients for each participant were transformed into three parameters: curvature at the vertex (**J**), center frequency (**K**), and maximum number of synapses per IHC (**L**). For **J-L**, the change in each parameter resulting from individualizing the CMAP relative to the value when the synaptogram was fit using a normal hearing (NH) pole function is shown. The change in value is plotted against the value from the synaptogram fit to a NH pole function. Dots indicate individual participants, colors indicate the evoked potentials that were used to predict synapse numbers, and the solid black line indicates no change. See Priors for Synapse Counts in Methods for details regarding the transformation of each coefficient into interpretable values.

### Impact of individualizing the CMAP with DPgrams

To assess whether individualizing the CMAP for each participant’s DPgram affected the predicted degree of deafferentation, synaptograms were regenerated for each participant under the assumption that all participants had the same normal hearing pole function. Each participant’s synaptogram was modeled by a parabola that is defined by three coefficients representing the curvature at the vertex, the center frequency (i.e., cochlear frequency of the vertex), and the maximum number of synapses per IHC. Notably, individualizing the CMAP using the DPgram had little impact on any of the synaptopgram coefficients generated by ABR_4_ (Figure 3J-L). In contrast, synaptograms generated by fits to EFR_4_ or both ABR_4_ and EFR_4_ were affected by individualizing the CMAP. This suggests that ABR wave I amplitude, in contrast to EFR magnitude, is a robust measure of auditory nerve function even in the presence of OHC dysfunction (up to a mild hearing loss).

### Validating synapse predictions using age

#### Proof of concept using mouse data

Since there is currently no means of directly validating predictions of cochlear deafferentation in living humans, many studies have turned to proxy measures such as performance on complex speech perception tasks (e.g., Garrett et al., 2025; Grant et al., 2020; Harris et al., 2021; Mepani et al., 2021).

However, post-mortem temporal bone data demonstrate that age is a key predictor of cochlear synapse counts (Wu et al., 2019). As a proof-of-concept, cochlear synapse counts from mice aged 10-104 weeks without noise exposure (Buran et al., 2026) were used to assess how well the number of synapses per IHC can predict mouse age. Although there was considerable variability in cochlear synapse numbers within age groups (e.g., 14 to 22 synapses/IHC for the youngest age group), there was an inverse correlation between synapse count and age (Figure 4A). A linear regression model was used to predict mouse age from synapse counts (e.g., 16 kHz, Figure 4B). Model performance was quantified as the root-mean-squared error (RMSE) of the difference in predicted vs. actual age (Figure 4C). Except for synapse counts at 5.6 kHz (the most apical region of the cochlea), synapse counts across cochlear frequency did a good job of predicting the age of each mouse (prediction error of 22.6-30.5 weeks). To provide a more direct parallel to our method of predicting age from the individualized synaptograms in humans, synapse count data from the mice were also fit to Equation 1 (shown as ***θ_i_*** in Figure 4C). The resulting mouse-specific synaptograms performed well at predicting the age of each mouse (prediction error of 22.2 weeks).

**Figure 4.**
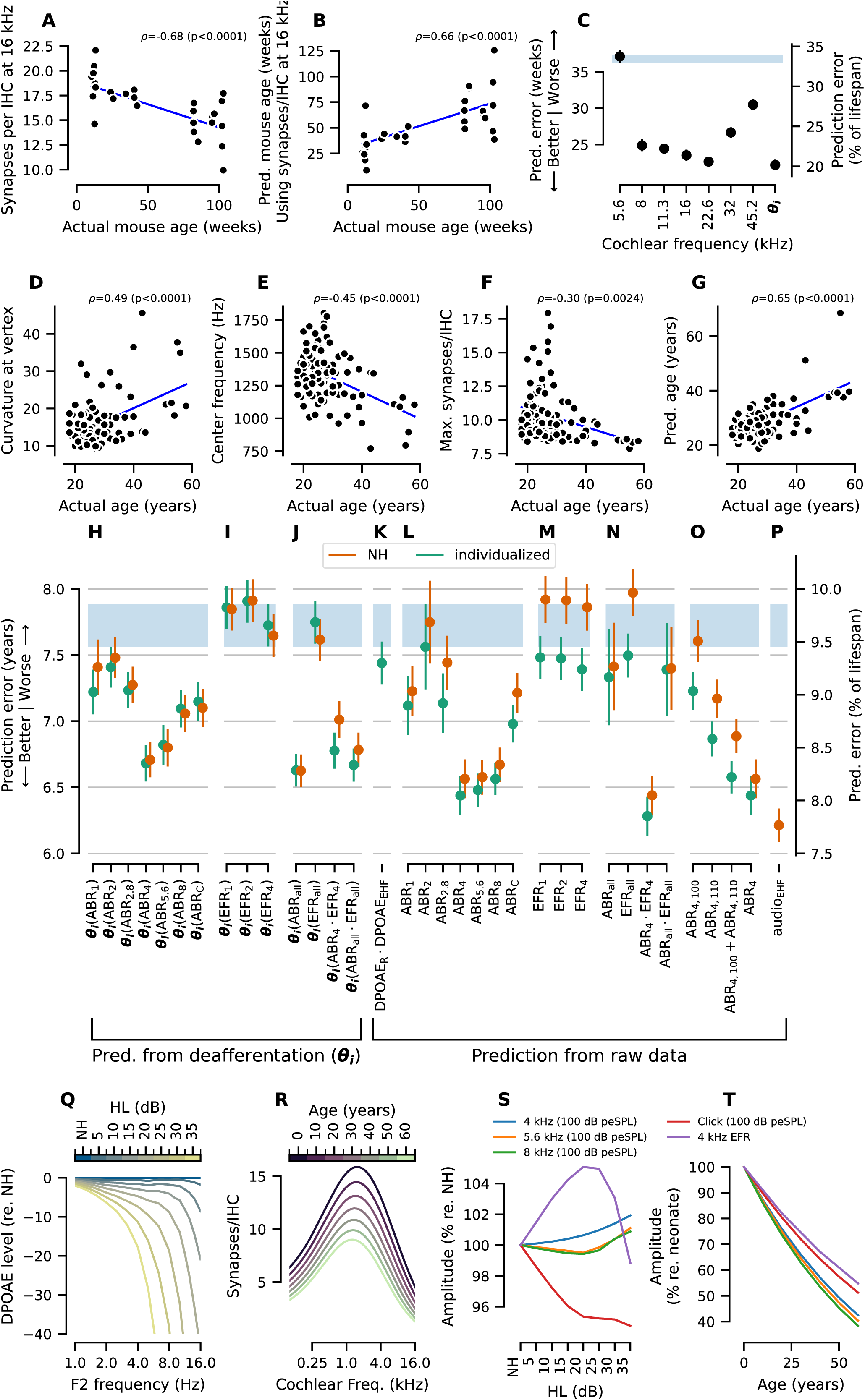
Comparison of linear regression models for their ability to predict age in mice or human participants. As a proof-of-concept, synapse count data collected from CBA/CaJ mice spanning a large range of ages (Buran et al. 2026) was reanalyzed. (**A**) Synapses per IHC at 16 kHz were inversely correlated with the age of the mouse. (**B**) Cross validation using a linear regression model shows that predicted synapse count at 16 kHz can be used to predict mouse age. (**C**) Prediction error for predicting mouse age from synapse counts for each cochlear frequency. To parallel the human analysis of synaptograms vs. age, synaptograms were also fit to the synapse count data for mouse. The resulting mouse-specific vector of three synaptogram coefficients (***θ_i_***) was then used to predict mouse age. The shaded blue region indicates baseline performance, as indicated by an intercept-only model in which the age is set to the average age of the mice in the sample. Data from 31 mice were included. Results that overlap with the blue region (e.g., 5.6 kHz) indicate that the model was not predictive of mouse age. The resulting prediction error, in weeks (left axis), is also translated to prediction error as a percentage of the median lifespan of a CBA/CaJ mouse (right axis, 110 weeks; Park et al., 2024). Better performance is indicated by smaller prediction error. For human participants, **D-F** shows the relationship between each coefficient of the synaptogram and participant age: curvature at the vertex (**D**), center frequency (**E**), and the maximum number of synapses per IHC (**F**). (**G**) Cross validation using a linear regression model incorporating the three synaptogram coefficients as predictors was used to predict participant age (y-axis). These predictions were compared with actual age (x-axis). The predicted age was set to the average across all repeats and folds. Solid regression lines in **D-G** in each plot show the best fit to the data. Markers indicate data from individual participants. Pearson’s correlation coefficient and p-value are provided above each plot. (**H-P**) Ability of linear regression models to predict participant age. Prediction error is reported as the root-mean-squared error (RMSE) of the difference between the participant’s actual versus predicted age. Models that used the synaptogram coefficients (***θ_i_***) to predict participant age (**H-J**) are grouped by the combination of evoked potentials used to generate the synaptograms: ABR wave I amplitude for a single frequency (**H**), EFR magnitude for a single frequency (**I**), or combination of ABR and/or EFR measurements at multiple frequencies (**J**). The stimuli used to generate the predictions are indicated as ***θ_i_***(*stimuli*). For all ABR models, both stimulus levels were used for generating the synapse predictions. For ABR_all_, the click ABR and all six ABR tone frequencies (1 through 8 kHz) were used for generating the synapse predictions. For EFR_all_, all three EFR carrier frequencies were used for generating the synapse predictions. Predictions were generated using a CMAP individualized for each participant’s DPgram (individualized; green points) or under the assumption that each participant had normal cochlear gain (NH; orange points). Models that used raw DPOAE or evoked potential measures to predict participant age (**K-O**) are grouped by measure: DPOAE (**K**), single ABR frequencies (**L**), single EFR frequencies (**M**), or combination of multiple ABR and/or EFR frequencies (**N**). (**O**) ABR models for a 4 kHz stimulus used either a single ABR level (ABR_4,100_ or ABR_4,110_), both ABR levels (ABR_4,100_ + ABR_4,110_), or both ABR levels including the interaction (ABR_4,100_ × ABR_4,110_) to predict participant age. (**P**) Model using extended high frequency audiometric data (average of pure tone detection thresholds ≥ 9 kHz; audio_EHF_). For panels **H-P**, the shaded blue region indicates baseline performance +/- standard error of the mean (SEM) for a model where the predicted age was based on sex only. Results that overlap with the blue region (e.g., NH ***θ_i_***(EFR_1_) in panel **I**) indicate that the model was not predictive of participant age. Better performance is indicated by smaller RMSE. Markers indicate the RMSE of the corresponding model averaged across all repeats and folds and error bars indicate the SEM. The resulting prediction error, in years (left axis), is also translated to prediction error as a percentage of the median human lifespan (right axis, 78.4 years; (Arias et al., 2025). (**Q-T**) Computational simulations of ABR wave I amplitude and EFR magnitude for different degrees of high frequency hearing loss and age-related deafferentation. (**Q**) High frequency hearing loss (HL) was modeled as a sloping loss that began at 1 kHz and reached the specified loss, in dB HL, at 8 kHz. The change in simulated DP level relative to simulated DP levels assuming normal hearing (NH) are shown for each hearing loss profile. (**R**) Simulated number of functional synapses per IHC with advancing age as estimated from the Wu et al. (2019) dataset. (**S**) Change in simulated ABR wave I amplitude or EFR magnitude with increasing high frequency HL relative to NH. In this simulation, it is assumed that no deafferentation occurs. (**T**) Change in simulated ABR wave I amplitude or EFR magnitude with increasing deafferentation using the synaptograms in **R**. In this simulation, it is assumed that there is no high frequency HL.

#### Predicting age from the individualized synaptograms

Consistent with expectations, each of the three coefficients defining the participant synaptograms (fit to ABR_4_) were correlated with participant age (Figure 4D-F). With advancing age, the predicted maximum number of synapses decreased, the center frequency shifted towards the apex, and the curvature at the vertex increased (i.e., the parabola became narrower).

All three coefficients were used in linear regressions models to predict the age of each participant (Figure 4G-O). Although the CMAP includes a correction for sex, all regression models included sex as a predictor (including the baseline model) to facilitate comparison with age predictions generated using the raw ABR, EFR, and/or DPOAE data, which may require a correction for sex. Thus, the baseline model (blue bar in Figure 4H-O) represents age predictions based on the average age for all males or all females in the sample, depending on the participant’s sex (as opposed to the average age of the full sample). Prediction error was then quantified for each synaptogram as generated by the individualized CMAP fit to various subsets of the evoked potential measurements (green markers in Figure 4H-J). To assess whether individualization of the CMAP led to improved age prediction performance, the models were refit using a CMAP with a normal hearing pole function (orange markers in Figure 4H-J). Out of all the ABR/EFR measurements or combinations of measurements used to generate synaptograms, synaptograms generated from ABR_all_ (regardless of individualization for the DPgram) performed the best (RMSE 6.63 SEM ± 0.12 years for both individualized and NH); however, ABR_all_ · EFR_all_ (individualized for the DPgram; RMSE 6.67 SEM ± 0.12 years) and ABR_4_ (regardless of individualization for the DPgram; individualized RMSE 6.68 SEM ± 0.14 years; NH RMSE 6.71 SEM ± 0.13 years) performed almost as well. Synaptograms generated from EFR measurements alone did not perform better than the baseline model (Figure 4I). Only ABR_1_, ABR_4_ · EFR_4_, and ABR_all_ · EFR_all_ demonstrated a benefit of individualizing the CMAP, with improvements in prediction error of 0.19 (± 0.07 SEM), 0.23 (± 0.05 SEM), and 0.12 (± 0.03 SEM) years, respectively.

#### Predicting age from raw physiological responses

As an alternative to the phenomenologically-inspired data reduction approach of combining multiple measures into three coefficients describing each participant’s synaptogram, simple linear regression models can be used to predict participant age based on one or more auditory physiological measurement (ABR, EFR, and DPOAE; Figure 4K-O). As for the synaptogram prediction models, the age prediction performance of the baseline model (average age for males/females in the sample; blue bar in Figure 4K-O) was plotted against the results for all other regression models. A regression model with only DPOAE_R_ and DPOAE_EHF_ as predictors (Figure 4K) provides a reference for comparing models that include evoked potentials as predictors. Due to the collinearity between evoked potentials and DPOAEs, the actual value of the regression coefficients (i.e., *β* in the equations) cannot be interpreted when more than one predictor is included in the model. However, the predictive power of these models is not affected by multicollinearity (Kutner et al., 2005).

With the exception of the ABR_2_ model, linear regression models incorporating raw ABR measurements performed better than the baseline model and the DPOAE-only model (Figure 4L). High frequency ABR models (ABR_4_, ABR_5.6_, and ABR_8_) outperformed low frequency ABR models (ABR_1_, ABR_2_, ABR_2.8_), and the click ABR model (ABR_c_). The best-performing model was ABR_4_ adjusted for DPOAEs (green markers in Figure 4L), which was able to predict participant age within 6.44 (±0.15 SEM) years. Excluding DPOAE_R_ and DPOAE_EHF_ from the model for ABR_4_ worsened the prediction error by 0.13 (±0.04 SEM) years (green markers vs orange markers in Figure 4L).

In contrast, models incorporating raw EFR measurements did not outperform the DPOAE-only model, even when adjusted for DPOAEs (green markers in Figure 4M). EFRs models that did not include an adjustment for DPOAEs (orange markers in Figure 4M) performed similarly to the baseline model, suggesting that the improved age prediction performance of the DPOAE-adjusted EFR models were driven largely by the inclusion of DPOAEs. Finally, a model that included both ABR_4_ and EFR_4_ and was adjusted for DPOAEs performed slightly better (RMSE 6.28 years, ±0.15 SEM) than the ABR_4_ model adjusted for DPOAEs, with an improvement in prediction error of 0.16 years (±0.04 SEM; Figure 4N). In contrast, including all of the ABR and EFR measurements in the same model resulted in age prediction performance that was only slightly better than the baseline model (Figure 4N).

Two stimulus levels were collected for each ABR frequency and both were included in the linear regression models shown in Figure 4L,N, along with an interaction between the two levels to account for potential compressive ABR wave I amplitude growth. Four additional linear regression models were evaluated to assess whether ABR_4_ predictions were driven by one or both stimulus levels (Figure 40). The regression model that included both intensity levels with an interaction between the levels (ABR_4_) and an adjustment for DPOAES performed the best. For the models that did not include an interaction between the stimulus levels, there was a larger difference in prediction error between the DPOAE-adjusted and the NH versions of the model than what was observed for ABR_4_.

Notably, the extended high frequency (EHF) audiometric thresholds (audio_EHF_) outperformed all other measures (Figure 4P) at predicting age, although it was only marginally better than the DPOAE-adjusted model that included both ABR_4_ and EFR_4_ with an improvement in prediction error of 0.07 years (±0.15 SEM).

#### Simulated impact of OHC dysfunction and age-related deafferentation on evoked potentials

DPOAEs are commonly used in statistical models to adjust ABR wave I amplitude for OHC dysfunction. However, the results of the current study, along with prior work in mouse (Buran et al., 2026), suggest that the ABR wave I amplitude in response to suprathreshold stimuli is less impacted by OHC dysfunction than previously expected. To verify these findings, computational simulations were used to assess the impact of both high frequency OHC dysfunction (as modeled by adjusting cochlear gain; Figure 4Q) and age-related deafferentation (Figure 4R) on ABR wave I amplitude and EFR magnitude. These simulations show that high frequency suprathreshold ABR wave I amplitude is less impacted by OHC dysfunction (Figure 4S) than the 4 kHz EFR or ABR wave I amplitude for a suprathreshold click stimulus. However, the differences across stimuli in the simulated impact of OHC dysfunction are small. For the greatest high frequency hearing loss (35 dB HL), there is a 1-2% increase in amplitude for high frequency toneburst ABR wave I amplitudes versus a 5% decrease in click ABR wave I amplitude and a 1 % decrease in the RAM EFR relative to normal hearing. Notably, for more moderate high frequency hearing loss (20 dB HL), the RAM EFR had a 5% increase in amplitude relative to normal hearing. In contrast, differences across stimuli in terms of the impacts of deafferentation are more considerable (Figure 4T), with a 58-62% decrease in magnitude for high frequency ABR tonebursts at 60 years (relative to a neonate) versus a 50% decrease for the click ABR and a 45% decrease for the 4 kHz RAM EFR.

#### Impacts of age-related declines in the endolymphatic potential

Changes in the endolymphatic potential (EP), which can affect both OHC and IHC function, may also contribute to age-related declines in auditory function. Although experimental reduction in EP leads to an acute decrease in active amplification, DPOAE levels can fully recover over time even if the cochlear microphonic remains reduced (Wang et al., 2019). Thus, the process of individualizing the CMAP using participant-specific DPgrams is expected to account for any EP-specific effects on OHC-based cochlear gain. EP-related changes in IHC function would not be reflected in temporal bone estimates of age-related IHC loss but could have age-related impacts on ABR and EFR measurements. Under the assumption that cochlear gain and tuning was preserved, the CMAP was used to model the effect of changes in EP on ABR wave I amplitude and EFR magnitude for 4 kHz stimuli (Figure 5A-B). Notably, EFR magnitude declined much more steeply than ABR wave I amplitude as EP was reduced (Figure 5B vs 5A), suggesting that the ABR is less impacted by changes in EP. Next, the CMAP was updated to support participant-specific predictions of EP (*EP*_i_) in addition to participant-specific synaptograms (***θ***_i_). Since decreases in both the EP and the synaptogram can lead to decreased ABR wave I amplitudes and EFR magnitudes, we fit four variants of the CMAP to ABR_4_ · EFR_4_: 1) a baseline-only model in which the only free parameter was the sex-specific scaling factor (baseline), 2) an EP-only model (***EP_i_***) in which the free parameters were the sex-specific scaling factor and *EP*_i_ - the synaptogram was fixed at that of a neonate, 3) a synaptogram-only model (***θ_i_***) in which the free parameters were the sex-specific scaling factor and ***θ_i_*** - the EP was fixed at 90 mV (***θ_i_***), and 4) an EP-synaptogram model (***EP_i_*** ⋅ 0_i_) in which the free parameters were the sex-specific scaling factor, *EP*_i_, and ***θ_i_***. The baseline-only model produced simulated ABR (Figure 5C) or EFR (Figure 5D) measurements in which the participant’s measurements were set to the average ABR or EFR measurements for the participant’s sex. Individualization of the CMAP using *EP*_i_ had a greater impact on simulated EFR magnitudes than simulated ABR magnitudes (Figure 5E-F), with a significant correlation between simulated and measured EFR magnitudes, but not ABR wave I amplitudes. Individualization using ***θ***_i_ resulted in significant correlations between simulated and measured ABR wave I amplitudes (Figure 5G), but not EFR magnitudes (Figure 5H). The ***EP_i_*** ⋅ ***θ***_i_ model performed as well as the ***θ***_i_ model in simulating participant-specific differences in ABR wave I amplitude (Figure 5I) and performed better than the ***θ***_i_ model in simulating participant-specific differences in EFR magnitude (Figure 5J).

**Figure 5.**
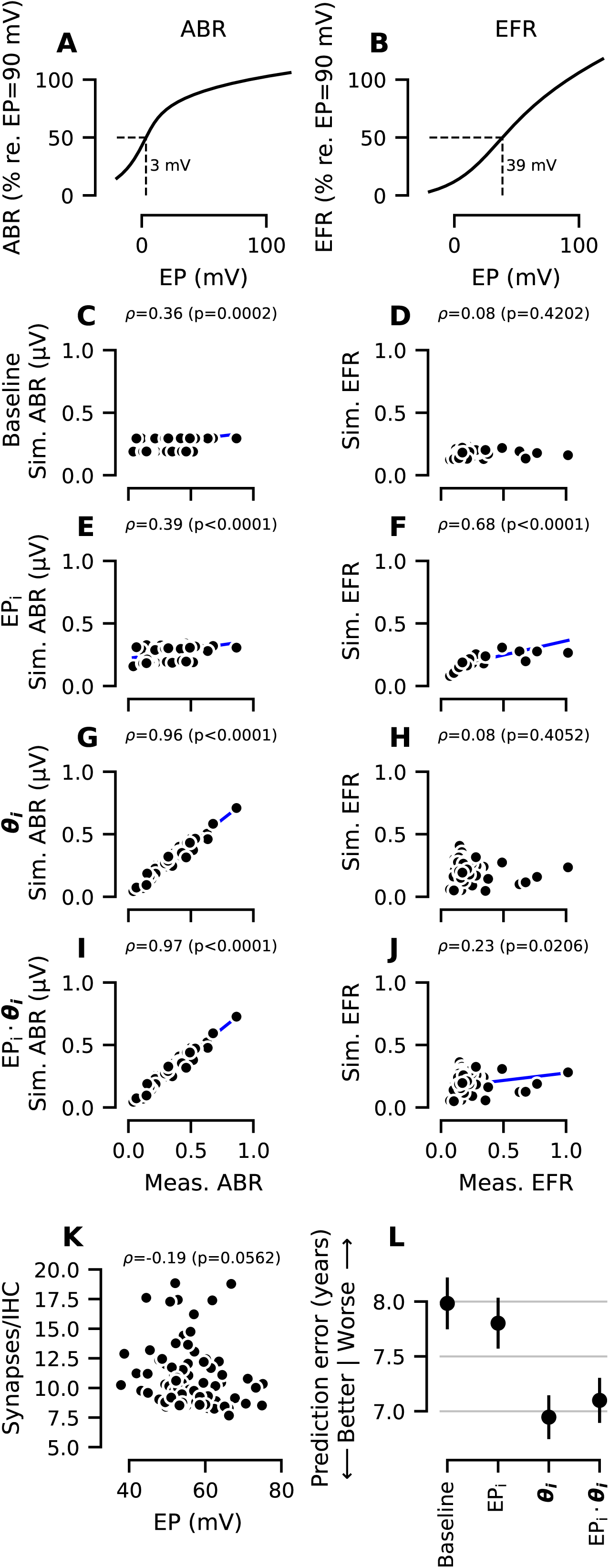
Age-related reductions in ABR wave I amplitude are better explained by cochlear deafferentation than age-related declines in endolymphatic potential. Modeled effect of endolymphatic potential (EP) on ABR wave I amplitude for 100 (green) and 110 (red) dB peSPL 4 kHz stimuli (A) and 4 kHz RAM EFR magnitude (B). The dashed line in A and B indicates the EP at which the evoked potential measure reaches 50% of the amplitude expected under normal, healthy EP conditions. Predictions of ABR wave I amplitude (C, E, G, I) and EFR magnitude (D, F, H, J) for a 4 kHz stimulus from models fit using ABR_4_ · EFR_4_. To assess the relative impact of EP vs. deafferentation (*θ*) on model predictions, four models were fit to the data: a baseline model including only sex as a predictor (C, D), a model including EP (EP_i_) as a free parameter (E, F), a model including deafferentation (*θ_i_*) as a free parameter (G, H), and a model including both EP and deafferentation as free parameters (*EP*_i_ ⋅ *θ_i_*) (I, J). For C-J, Pearson’s correlation coefficients and p-values are shown above each plot. A solid regression line is shown only for p < 0.05. (K) For the model incorporating both deafferentation and EP (E, J), there was no significant correlation between predicted deafferentation and predicted EP, suggesting that the model is able to separate these two parameters. (L) Ability of each model to predict participant age as measured by the RMSE of the prediction error.

Since either a reduction in the EP or deafferentation can be used to simulate decreased evoked potentials, a correlation between *EP*_i_ and ***θ***_i_ would be expected if the model was unable to separate these parameters (i.e., if the model predicted a high EP for a given participant, it would decrease the synaptogram to compensate); however, these values were not significantly correlated (Figure 5K), suggesting that the model is able to separate the effects of these two parameters on evoked potentials. Next, we tested how well *EP*_i_ and ***θ***_i_ could predict age (Figure 5L). The baseline-only model, in which the predicted age of each participant was set to the average age for the participant’s sex, is shown for reference. The *EP*_i_ model performed similarly to the baseline model, while both the ***θ***_i_ model and the ***EP_i_*** ⋅ ***θ***_i_ model performed better than the baseline-only model at predicting participant age.

## Discussion

In a sample of 102 adults with relatively good hearing and a wide range of DPOAE, ABR, and EFR responses, a CMAP was used with Bayesian regression to predict cochlear deafferentation, represented as the frequency-specific number of functional auditory nerve fibers per IHC (synaptogram), for each participant. The CMAP can combine multiple DPOAE, ABR, and EFR measurements in response to different stimuli, thereby reducing a heterogenous set of measurements to a single synaptogram. This study expanded on Buran et al. (2022), where the CMAP was used with ABR wave I amplitude at multiple frequencies and levels to predict synaptograms in individuals with normal hearing thresholds, by 1) incorporating EFR measurements, 2) including participants with up to a mild sensorineural hearing loss, allowing for better assessment of the cochlear gain individualization, 3) incorporating extended high EHF DPOAEs, 4) collecting a broader range of ABR frequencies similar to animal studies of synaptopathy, and 5) incorporating the full shape of the synaptogram into comparisons with age rather than using only the coefficient representing the maximum number of functional auditory nerve fibers per IHC. Since the models did not include participant age as a predictor, the models are a measure of how well the incorporated coefficients can predict age, a key risk factor for cochlear deafferentation. The CMAP was also used to simulate the effect of the EP on the evoked potentials to determine whether age-related reductions in EP could offer an alternate explanation for the observed decline in the evoked potentials with age.

### Mouse data validates the ability of synapse counts to accurately predict age

Reanalysis of a subset (31 mice) of data from a study that histologically quantified cochlear synapses in mice encompassing a large age span (Buran et al., 2026) demonstrates that, despite variability in synapse counts across individual mice (14-22 synapses per IHC in the youngest mice), synapse counts can be used to predict mouse age significantly better than chance (Figure 4C). The observed variability in synapse counts among mice of the same age (Figure 4A) raises the possibility that much of the observed inter-subject variability in ABR wave 1/I amplitude seen in animals/humans may be due to variability in the actual degree of cochlear deafferentation across subjects rather than measurement error due to anatomical variation.

### High frequency ABR wave I amplitudes performed best at predicting participant age

Linear regression models incorporating either synaptograms generated from ABR_4_, ABR_all_, or ABR_all_ · EFR_all_; high frequency raw ABR wave I amplitudes (ABR_4_, ABR_5.6_, and ABR_8_); or raw ABR_4_ · EFR_4_ measurements performed best at predicting participant age. Combining multiple evoked potential measurements, either as independent predictors in a linear regression model or when predicting the synaptogram using the CMAP, generally did not offer large improvements in age prediction over single ABR_4_ measurements. While a prediction error of 6 to 6.5 years may not seem notably different from the baseline error of ∼7.75 years (Figure 4H-P), this represents a significant improvement in performance. For example, we can also look at the correlation between single measures and age. Raw ABR wave I amplitudes for a 4 kHz, 110 dB peSPL tone pip were inversely correlated with age (r=-0.51, p < 0.0001) and had an age prediction error of 7.2 years (ABR_4,110_; orange marker in Figure 4O), EHF audiometric thresholds were correlated with age (r=0.63, p < 0.0001) and had an age prediction error of 6.2 years (audio_EHF_; orange marker in Figure 4P), and the EFR magnitude in response to a 4 kHz RAM EFR tone (EFR4) was not correlated with age (r=-0.12, p=0.25) and had an age prediction error of 7.9 years (EFR_4_; orange marker in Figure 4M).

Incorporating two stimulus levels in the ABR_4_ models, particularly when an interaction between the two levels was also included, resulted in better age-prediction performance than for ABR_4_ models with only a single stimulus level (Figure 4O). This suggests that incorporating more than one intensity level may provide information about ABR wave I amplitude growth that can improve estimates of deafferentation from ABR measurements, consistent with previous studies (Garrett & Verhulst, 2019; Johannesen et al., 2019). The results of Buran et al. (2026) also suggested that ABR wave 1 amplitude growth may be helpful for predicting synapse numbers in mice.

Despite reported age-related declines in SAM (Bramhall et al., 2023) and RAM (Van Der Biest et al., 2023) EFR magnitude, RAM EFR was not a good predictor of age in this study. One possible explanation for the RAM EFR’s poor age prediction ability is that cochlear deafferentation leads to central gain in the auditory brainstem (Chambers et al., 2016; Salvi et al., 2017), which may confound interpretation of the RAM EFR since the auditory brainstem contributes to the EFR response at the 110 Hz modulation frequency used in this study (Kuwada et al., 2002). In Buran et al. (2026), the RAM EFR modulated at 110 Hz performed more poorly than ABR wave 1 amplitude at predicting synapse counts in mice, consistent with the results of the current study. Another possible explanation is that the relative performance of the ABR and the RAM EFR depends on the presentation level. The toneburst ABRs were presented at 100 and 110 dB peSPL, whereas the RAM EFR was presented at 70 dB SPL. The RAM EFR may predict age better at higher presentation levels where it will be less impacted by OHC dysfunction; however, such levels may be difficult to tolerate by participants.

Notably, the model that included EHF audiometric thresholds (audio_EHF_) outperformed all other models at predicting age. It is generally assumed that synaptopathy and IHC loss will not impact pure tone detection thresholds until the degree of deafferentation becomes extreme (Chambers et al., 2016; Lobarinas et al., 2013). However, Lobarinas et al. observed a threshold shift of approximately 20 dB when roughly 75% of IHCs were lost without any OHC loss. Histological data from human temporal bones shows precipitous steeply sloping losses of IHCs in the EHF region of the cochlea, with greater than 50% IHC loss for adults younger than 60 years (Wu et al., 2019). Although the ANF fiber data in the EHFs is sparse in the Wu et al. dataset, it suggests that EHF synapse numbers are low even in neonates (4-5 per IHC), with approximately 0.3 synapses/IHC lost per decade, equating to approximately 40% EHF ANF loss by age 60 years. Age-related EHF deafferentation due to IHC and synapse loss may contribute to elevated EHF audiometric thresholds. Hauser et al. (2025) showed that careful calibration of DPOAE stimuli improved the correlation between measured EHF DPOAE levels and EHF pure tone thresholds. However, the best correlation between these was approximately -0.5, while the correlation between EHF audiometric thresholds and age was 0.852, indicating that factors other than OHC function likely contribute to this correlation. Taken together, these findings suggest that age-related declines in EHF audiometric thresholds reflect the combined impacts of OHC dysfunction and deafferentation.

### Including DPOAEs had limited effect on the age-prediction ability of the best performing models

In terms of predicting age, there was no clear benefit in the best performing models of individualizing the synaptogram based on DPOAEs or including DPOAEs the raw data models. This is consistent with the results of Buran et al. (2026) where the ability to predict synapse counts in mice (which had a large range of auditory thresholds) with ABR wave 1 amplitude measurements was similar regardless of whether an adjustment for DPOAEs was used. This suggests that, at least for individuals with relatively good hearing (no more than a mild hearing loss), high frequency suprathreshold ABR wave I amplitudes can be used as an indicator of cochlear deafferentation without adjusting for OHC function.

This may be a bit surprising given the results of Verhulst et al. (2016), which used computational modeling to show that ABR wave I amplitude (for a click stimulus) is impacted by high frequency cochlear gain loss. However, additional simulations (Figure 4Q-T) indicate that the high frequency toneburst ABR stimuli used in this study are more robust to OHC dysfunction than the click ABR, which is often used in studies of human synaptopathy. These simulations also show a much greater impact of simulated synaptopathy on ABR wave I amplitude and EFR magnitude than the effect of high frequency hearing loss (Verhulst et al. 2016; Figure 4S). While there are subtle differences across the literature in terms of the relative impacts of synaptopathy and OHC dysfunction on ABR and EFR measures, this is likely due to the specific stimuli design and assumptions regarding the tonotopic distribution of ANFs and how ANFs are lost as a function of SR.

### Impact of endolymphatic potential on predicted deafferentation

Transient reductions in the EP have been shown to lead to reduced active amplification by OHCs; however, the cochlea adapts within hours to sustained reductions in the EP, with DPOAEs recovering to normal levels within several hours (Wang et al., 2019). It is assumed that the resulting drop in the EP can also impact IHC receptor potentials, leading to a decline in auditory nerve activity. The specific effect of a decline in the EP on IHC responses cannot easily be separated from the loss of active amplification by OHCs in vivo. However, our computational modeling suggests that while both ABR wave I amplitude and EFR magnitude can be affected by declines in the EP, the impacts on ABR wave I amplitude are more modest. In the current analysis, computational modeling in which the EP was allowed to vary across participants improved model predictions for the EFR but not for the ABR, suggesting that the EFR may offer more insight into the status of the EP. Models incorporating the EP as a free parameter did not provide any additional improvement in the ability to predict participant age, suggesting that deafferentation is a more likely explanation for the observed age-related decline in evoked potentials within the sample age range (up to 58 years). Indeed, prominent atrophy of the stria vascularis is a late-onset pathology primarily occurring in individuals over 60 years of age (Lang et al., 2023; Takahashi, 1971). While metabolic damage may represent a major contributor to auditory nerve activity in older cohorts with more severe hearing loss (Vaden et al., 2022), cochlear deafferentation appears to be the primary driver in the current young to middle-aged population with relatively intact cochlear gain.

### Utility of using the CMAP to predict synapse counts

Although the age prediction results suggest that using the CMAP may not result in better prediction of deafferentation than using raw ABR measurements, the CMAP offers a biophysically-inspired approach to inferring peripheral auditory function from measurements of otoacoustic emissions and evoked potentials. Although models using raw ABR and DPOAE measurements can outperform the CMAP in terms of predicting participant age, the high collinearity of the individual raw ABR and DPOAE measurements mean that the coefficients of these models cannot be translated into meaningful values and, thus, have limited applicability to future human studies of cochlear deafferentation where one might want predict deafferentation in individual participants. In contrast, once the neural network coefficients (for the DPgram) and the sex-specific and stimuli-specific scaling factors of the Bayesian regression have been determined, this approach can be used to predict deafferentation in individual participants.

Using the CMAP to predict deafferentation in individual participants also allows for data to be combined in a meta-analysis across studies that collected different ABR or EFR stimuli. For example, deafferentation could be predicted for all the participants in human studies that investigated the relationship between deafferentation and tinnitus even though the studies used different ABR and EFR measurements to estimate the degree of deafferentation. Synaptograms could then be evaluated for relationships with tinnitus across the participants for all the studies, resulting in substantially increased power.

In addition, the CMAP approach is more robust to overfitting than raw measurements when working with multiple ABR and/or EFR measurements collected in response to different stimuli. For example, the age-prediction model that included the full set of raw ABR and EFR measurements (ABR_all_ · EFR_all_) did not perform much better than baseline, likely due to overfitting. The large number of parameters relative to the size of the sample can result in the model having greater difficulty recognizing meaningful patterns (Hastie et al., 2009). While there are statistical techniques that can be used to address overfitting, these techniques can make it more challenging to interpret the results and are sometimes unstable with correlated predictors. A simpler, more interpretable approach is to analyze each predictor independently against the outcome; however, this leads to decreased statistical power due to the necessity of correcting for multiple comparisons. By defining structured relationships between DPOAEs, ABRs, and EFRs, the CMAP offers a simple approach to address the collinearity issue by pinpointing the theoretical source of the individual differences in ABR or EFR measurements – the synaptograms. The utility of using the CMAP when incorporating multiple ABR and EFR measurements is demonstrated by the superior ability of models that included synaptograms generated from all the ABR and EFR measurements to predict age compared to models that included all the raw ABR and EFR measurements (Figure 4J versus 4N).

### Limitations

Although the synaptogram nominally describes the number of synapses per IHC, it should be more generally thought of as a measure of cochlear deafferentation that reflects the number of functional auditory nerve fibers contributing to the auditory evoked potentials. There are many potential sources of deafferentation including synaptopathy, SGC loss, IHC damage/loss, and demyelination. The synaptogram is unable to differentiate between these different sources of deafferentation. Although the perceptual impacts of different sources of deafferentation are expected to be very similar, more precise differential diagnostics may be required in the future to identify appropriate therapeutic targets (e.g., IHC regeneration vs. synaptic regeneration).

Due to our inability to quantify cochlear synapse numbers in living humans, age (a known risk factor for cochlear synaptopathy) was used as a proxy for deafferentation in this analysis. Although age has been used in other studies for comparing different physiological indicators of deafferentation (e.g., Bharadwaj et al., 2022; Carcagno & Plack, 2020) and is consistent with observations from human temporal bones (Wu et al. 2019, 2021), it remains a crude proxy. There are other age-related factors that may contribute to changes in ABR wave I amplitude and EFR magnitude such as reductions in EP and demyelination. Computational simulations suggest that the EP cannot explain participant-specific differences in ABR wave I amplitude and that age prediction models incorporating participant-specific estimates of cochlear deafferentation significantly outperformed models that only incorporated participant-specific estimates of EP (Figure 5L). However, incorporating participant-specific estimates of EP notably improved estimates of the EFR (e.g., Figure 5F and 5J), suggesting a potential role for the EFR in a differential diagnosis of EP-mediated changes in IHC function versus other sources of deafferentation.

The limited utility of the EFR for predicting age stands in contrast to earlier empirical findings demonstrating an age-related decline in EFR magnitude (Van Der Biest et al., 2023), as well as computational models predicting high sensitivity of the RAM EFR to synaptopathy (Vasilkov et al., 2021). This unexpected divergence likely reflects the multi-generator nature of the EFR, which receives contributions from cortical, midbrain, and auditory nerve sources. Because our computational framework does not simulate central gain, compensatory plasticity along the central auditory pathway following peripheral deafferentation may mask peripheral synapse loss, thereby reconciling our empirical findings with model predictions.

This study did not measure head size, which is known to be inversely correlated with ABR wave I amplitude (Mitchell et al. 1989; Trune et al. 1987); however, head size would not be expected to covary with age and thus should not impact the overall conclusion. Future work incorporating head size as a parameter in the CMAP or a predictor in general linear regression models could potentially strengthen the relationships reported in this study.

Since there are known sex-related differences in ABR wave I amplitude, sex was treated as a scaling factor in the CMAP. We chose not to attempt to model the underlying biological basis of these sex-related differences since there is no definitive description of the underlying source of this variation. Even if this scaling factor is replaced by a biophysical model, the ultimate outcome of a biophysical model would be equivalent to the sex-related scaling factor.

## Conclusions

Both estimates of cochlear deafferentation generated using the CMAP and raw ABR wave I amplitude measurements (for a 4 kHz toneburst) performed well at predicting age, partially validating the ability of the CMAP to accurately predict deafferentation from ABR wave I amplitude data. This suggests that the CMAP can be used in the future to predict the degree of deafferentation in individual human participants based on their ABR measurements, a task that is not currently possible using raw ABR measurements due to our inability to determine accurate coefficients for predicting deafferentation from ABR wave I amplitudes. Computational simulations indicated that while both the ABR and EFR can be impacted by reductions in the EP after correcting for OHC function, deafferentation is a more plausible explanation of the age-related decline in ABR and EFR measurements observed in the current study than changes to the EP.

Of the deafferentation predictions and raw ABR and EFR measurements that were evaluated, deafferentation predictions generated from ABR wave I amplitudes for a 4 kHz toneburst and raw ABR wave I amplitudes for a 4 kHz toneburst were the best single measurement predictors of age. This suggests that 4 kHz ABR wave I amplitudes are better than other evoked potential measures at predicting age-related cochlear deafferentation. Combining raw 4 kHz ABR wave I amplitude and RAM EFR measurements may slightly improve the ability to detect cochlear deafferentation, but the additional benefit may not outweigh the extra time required to collect both measurements. Accounting for DPOAEs provided limited benefit for the synaptopgram and raw ABR_4_ regression models in terms of improving the ability to predict age. This suggests that, in individuals with up to a mild hearing loss, it may not be necessary to adjust high frequency ABR wave I amplitudes for DPOAEs when estimating cochlear deafferentation.

## Data Availability

All data produced in the present study are available upon reasonable request to the corresponding author.

## Acknowledgements

This work was supported by the National Institutes of Health, National Institute on Deafness and Other Communication Disorders - Award #R01DC020423 (to N.F.B). and by resources and facilities at the VA National Center for Rehabilitative Auditory Research (NCRAR) [Center Award #C2361C/I50 RX002361] at the VA Portland Health Care System in Portland, OR. The opinions and assertions presented are private views of the authors and are not to be construed as official or as necessarily reflecting the views of the National Institutes of Health or the Department of Veterans Affairs.

## Author contribution statement

BNB, NFB, and SV designed the experiments. SDK, AEH, NKW, and HAS performed the experiments. BNB and MT provided computational methods. BNB performed the analysis. BNB and NFB wrote the paper. All authors contributed edits to the manuscript. NFB provided funding for the study.

---

**Supplemental Data Figure 1.**
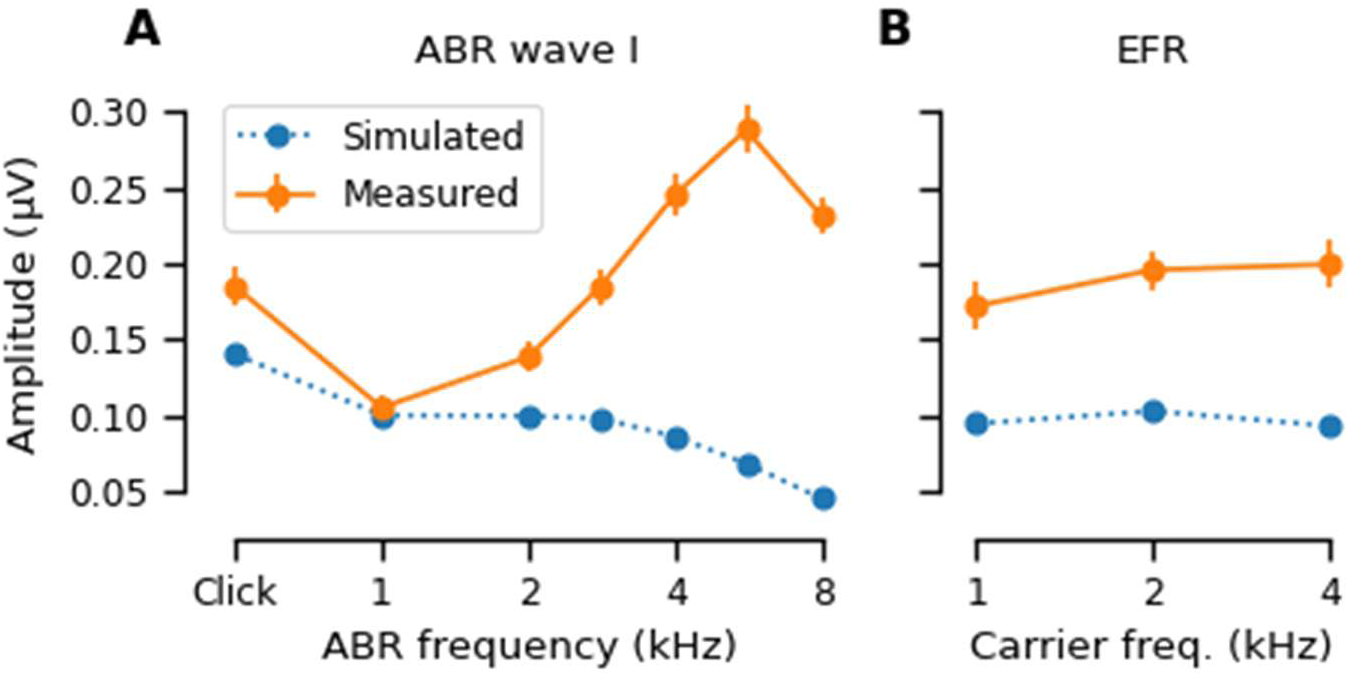
Differences between simulated and measured evoked potentials. Simulated amplitudes (blue dashed line) for ABR wave I (A) and EFR (B) were generated with the CMAP under the assumption that participants had normal-hearing pole functions with a normal complement of auditory nerve fiber synapses. Measured amplitudes (orange solid line) were calculated as the mean measurements collected from the non-Veteran participants in the sample. Error bars indicate standard error of the mean (SEM).

## Notes

### Competing Interest Statement

The authors have declared no competing interest.

### Author Declarations

The IRB of the Veterans Affairs Portland Health Care System (VAPORHCS) gave ethical approval for this work.

### Summary of Updates

Revision has been updated to improve the DP-gram prediction model, significantly minimizing the errors seen at high F2 frequencies. Additional analyses of the endocochlear potential have been added. Discussion has greatly been expanded. To make room for these additions, the analysis of Veteran status has been removed.

## References

1. Arias, E., Xu, J., & Kochanek, K. (2025). United States Life Tables, 2023. Retrieved June 8 from 10.15620/cdc/174591

2. Bharadwaj, H. M., Hustedt-Mai, A. R., Ginsberg, H. M., Dougherty, K. M., Muthaiah, V. P. K., Hagedorn, A., Simpson, J. M., & Heinz, M. G. (2022). Cross-species experiments reveal widespread cochlear neural damage in normal hearing. Commun Biol, 5(1), 733. 10.1038/s42003-022-03691-4

3. Bramhall, N. F. (2021). Use of the auditory brainstem response for assessment of cochlear synaptopathy in humans. J Acoust Soc Am, 150(6), 4440. 10.1121/10.0007484

4. Bramhall, N. F., & McMillan, G. P. (2024). Perceptual Consequences of Cochlear Deafferentation in Humans. Trends Hear, 28, 23312165241239541. 10.1177/23312165241239541

5. Bramhall, N. F., McMillan, G. P., & Kampel, S. D. (2021). Envelope following response measurements in young veterans are consistent with noise-induced cochlear synaptopathy. Hear Res, 408, 108310. 10.1016/j.heares.2021.108310

6. Bramhall, N. F., Theodoroff, S. M., McMillan, G. P., Kampel, S. D., & Buran, B. N. (2023). Associations Between Physiological Correlates of Cochlear Synaptopathy and Tinnitus in a Veteran Population. J Speech Lang Hear Res, 66(11), 4635–4652. 10.1044/2023_JSLHR-23-00234

7. Buran, B. N. (2015). Auditory-wave-analysis: v1.1. Retrieved April 14, 2017 from http://zenodo.org/record/17365#.VoMCrjbUi70

8. Buran, B. N., Elkins, S., He, W., & Bramhall, N. F. (2025). Predicting cochlear synaptopathy in mice with varying degrees of outer hair cell dysfunction using auditory evoked potentials. bioRxiv, 2025.2005.2009.653157. 10.1101/2025.05.09.653157

9. Buran, B. N., Elkins, S., He, W., Verhulst, S., & Bramhall, N. F. (2026). Predicting Cochlear Synaptopathy in Mice with Varying Degrees of Outer Hair Cell Dysfunction Using Auditory Evoked Potentials. J Assoc Res Otolaryngol, 27(1), 61–81. 10.1007/s10162-025-01015-x

10. Buran, B. N., McMillan, G. P., Keshishzadeh, S., Verhulst, S., & Bramhall, N. F. (2022). Predicting synapse counts in living humans by combining computational models with auditory physiology. J Acoust Soc Am, 151(1), 561. 10.1121/10.0009238

11. Burman, P. (1989). A comparative study of ordinary cross-validation, v-fold cross-validation and the repeated learning-testing methods. Biometrika, 76(3), 503–514. 10.1093/biomet/76.3.503

12. Carcagno, S., & Plack, C. J. (2020). Effects of age on electrophysiological measures of cochlear synaptopathy in humans. Hear Res, 396, 108068. 10.1016/j.heares.2020.108068

13. Chambers, A. R., Resnik, J., Yuan, Y., Whitton, J. P., Edge, A. S., Liberman, M. C., & Polley, D. B. (2016). Central Gain Restores Auditory Processing following Near-Complete Cochlear Denervation. Neuron, 89(4), 867–879. 10.1016/j.neuron.2015.12.041

14. Dolphin, W. F., & Mountain, D. C. (1992). The envelope following response: scalp potentials elicited in the Mongolian gerbil using sinusoidally AM acoustic signals. Hear Res, 58(1), 70–78. 10.1016/0378-5955(92)90010-k

15. Encina-Llamas, G., Harte, J. M., Dau, T., Shinn-Cunningham, B., & Epp, B. (2019). Investigating the Effect of Cochlear Synaptopathy on Envelope Following Responses Using a Model of the Auditory Nerve. J Assoc Res Otolaryngol, 20(4), 363–382. 10.1007/s10162-019-00721-7

16. Furman, A. C., Kujawa, S. G., & Liberman, M. C. (2013). Noise-induced cochlear neuropathy is selective for fibers with low spontaneous rates. J Neurophysiol, 110(3), 577–586. 10.1152/jn.00164.2013

17. Garrett, M., Vasilkov, V., Mauermann, M., Devolder, P., Wilson, J. L., Gonzales, L., Henry, K. S., & Verhulst, S. (2025). Deciphering Compromised Speech-in-Noise Intelligibility in Older Listeners: The Role of Cochlear Synaptopathy. eNeuro, 12(2). 10.1523/ENEURO.0182-24.2024

18. Garrett, M., & Verhulst, S. (2019). Applicability of subcortical EEG metrics of synaptopathy to older listeners with impaired audiograms. Hear Res, 380, 150–165. 10.1016/j.heares.2019.07.001

19. Grant, K. J., Mepani, A. M., Wu, P., Hancock, K. E., de Gruttola, V., Liberman, M. C., & Maison, S. F. (2020). Electrophysiological markers of cochlear function correlate with hearing-in-noise performance among audiometrically normal subjects. J Neurophysiol, 124(2), 418–431. 10.1152/jn.00016.2020

20. Gratton, M. A., Schmiedt, R. A., & Schulte, B. A. (1996). Age-related decreases in endocochlear potential are associated with vascular abnormalities in the stria vascularis. Hear Res, 94(1-2), 116–124. 10.1016/0378-5955(96)00011-1

21. Greenwood, D. D. (1990). A cochlear frequency-position function for several species--29 years later. J Acoust Soc Am, 87(6), 2592–2605. 10.1121/1.399052

22. Harris, K. C., Ahlstrom, J. B., Dias, J. W., Kerouac, L. B., McClaskey, C. M., Dubno, J. R., & Eckert, M. A. (2021). Neural Presbyacusis in Humans Inferred from Age-Related Differences in Auditory Nerve Function and Structure. J Neurosci, 41(50), 10293–10304. 10.1523/JNEUROSCI.1747-21.2021

23. Hashimoto, I., Ishiyama, Y., Yoshimoto, T., & Nemoto, S. (1981). Brain-stem auditory-evoked potentials recorded directly from human brain-stem and thalamus. Brain, 104(Pt 4), 841–859.

24. Hastie, T., Tibshirani, R., & Friedman, J. (2009). The Elements of Statistical Learning: Data Mining, Inference, and Prediction (2nd ed.). Springer-Verlag.

25. Hauser, S. N., Hustedt-Mai, A. R., Wichlinski, A., & Bharadwaj, H. M. (2025). The relationship between distortion product otoacoustic emissions and audiometric thresholds in the extended high-frequency range. J Acoust Soc Am, 157(3), 1889–1898. 10.1121/10.0036143

26. Herdman, A. T., Lins, O., Van Roon, P., Stapells, D. R., Scherg, M., & Picton, T. W. (2002). Intracerebral sources of human auditory steady-state responses. Brain Topogr, 15(2), 69–86. 10.1023/a:1021470822922

27. Hoffman, M. D., & Gelman, A. (2014). The No-U-Turn Sampler: Adaptively Setting Path Lengths in Hamiltonian Monte Carlo. J Mach Learn Res, 15, 1593–1623.

28. Johannesen, P. T., Buzo, B. C., & Lopez-Poveda, E. A. (2019). Evidence for age-related cochlear synaptopathy in humans unconnected to speech-in-noise intelligibility deficits. Hear Res, 374, 35–48.

29. Keshishzadeh, S., & Verhulst, S. (2021). Individualized Cochlear Models Based on Distortion Product Otoacoustic Emissions. Conf Proc IEEE Eng Med Biol Soc(2021), 403–407. 10.1109/EMBC46164.2021.9629808

30. Kiren, T., Aoyagi, M., Furuse, H., & Koike, Y. (1994). An experimental study on the generator of amplitude-modulation following response. Acta Otolaryngol Suppl, 511, 28–33.

31. Konrad-Martin, D., Poling, G. L., Dreisbach, L. E., Reavis, K. M., McMillan, G. P., Lapsley Miller, J. A., & Marshall, L. (2016). Serial Monitoring of Otoacoustic Emissions in Clinical Trials. Otol Neurotol, 37(8), e286–294. 10.1097/MAO.0000000000001134

32. Krizman, J., Skoe, E., & Kraus, N. (2012). Sex differences in auditory subcortical function. Clin Neurophysiol, 123(3), 590–597. 10.1016/j.clinph.2011.07.037

33. Kujawa, S. G., & Liberman, M. C. (2009). Adding insult to injury: cochlear nerve degeneration after “temporary” noise-induced hearing loss. J Neurosci, 29(45), 14077–14085. 10.1523/JNEUROSCI.2845-09.2009

34. Kutner, M. H., Nachtsheim, C. J., Neter, J., & Li, W. (2005). Applied Linear Statistical Models (5th ed.). McGraw-Hill Irwin.

35. Kuwada, S., Anderson, J. S., Batra, R., Fitzpatrick, D. C., Teissier, N., & D’Angelo, W. R. (2002). Sources of the scalp-recorded amplitude-modulation following response. J Am Acad Audiol, 13(4), 188–204.

36. Lang, H., Noble, K. V., Barth, J. L., Rumschlag, J. A., Jenkins, T. R., Storm, S. L., Eckert, M. A., Dubno, J. R., & Schulte, B. A. (2023). The Stria Vascularis in Mice and Humans Is an Early Site of Age-Related Cochlear Degeneration, Macrophage Dysfunction, and Inflammation. J Neurosci, 43(27), 5057– 5075. 10.1523/JNEUROSCI.2234-22.2023

37. Liberman, M. C. (1978). Auditory-nerve response from cats raised in a low-noise chamber. J Acoust Soc Am, 63(2), 442–455.

38. Lobarinas, E., Salvi, R., & Ding, D. (2013). Insensitivity of the audiogram to carboplatin induced inner hair cell loss in chinchillas. Hear Res, 302, 113–120. 10.1016/j.heares.2013.03.012

39. Mepani, A. M., Verhulst, S., Hancock, K. E., Garrett, M., Vasilkov, V., Bennett, K., de Gruttola, V., Liberman, M. C., & Maison, S. F. (2021). Envelope following responses predict speech-in-noise performance in normal-hearing listeners. J Neurophysiol, 125(4), 1213–1222. 10.1152/jn.00620.2020

40. Moller, A. R., & Jannetta, P. J. (1981). Compound action potentials recorded intracranially from the auditory nerve in man. Exp Neurol, 74(3), 862–874.

41. Neely, S., & Liu, Z. (1993). EMAV: Otoacoustic emission averager. In *Tech Memo* No. 17. Boys Town National Research Hospital Omaha.

42. Park, C. R., Willott, J. F., & Walton, J. P. (2024). Age-related changes of auditory sensitivity across the life span of CBA/CaJ mice. Hear Res, 441, 108921. 10.1016/j.heares.2023.108921

43. Parthasarathy, A., & Kujawa, S. G. (2018). Synaptopathy in the Aging Cochlea: Characterizing Early-Neural Deficits in Auditory Temporal Envelope Processing. J Neurosci, 38(32), 7108–7119. 10.1523/JNEUROSCI.3240-17.2018

44. Paul, B. T., Waheed, S., Bruce, I. C., & Roberts, L. E. (2017). Subcortical amplitude modulation encoding deficits suggest evidence of cochlear synaptopathy in normal-hearing 18-19 year olds with higher lifetime noise exposure. J Acoust Soc Am, 142(5), EL434. 10.1121/1.5009603

45. Purcell, D. W., John, S. M., Schneider, B. A., & Picton, T. W. (2004). Human temporal auditory acuity as assessed by envelope following responses. J Acoust Soc Am, 116(6), 3581–3593. 10.1121/1.1798354

46. Salvatier, J., Wiecki, T., & Fonnesbeck, C. (2016). Probabilistic programming in Python using PyMC3. PeerJ Comput Sci, 2, e55.

47. Salvi, R., Sun, W., Ding, D., Chen, G.-D., Lobarinas, E., Wang, J., Radziwon, K., & Auerbach, B. D. (2017). Inner hair cell loss disrupts hearing and cochlear function leading to sensory deprivation and enhanced central auditory gain. Front Neurosci, 10, 621. 10.3389/fnins.2016.00621

48. Schmiedt, R. A., Mills, J. H., & Boettcher, F. A. (1996). Age-related loss of activity of auditory-nerve fibers. J Neurophysiol, 76(4), 2799–2803. 10.1152/jn.1996.76.4.2799

49. Schulte, B. A., & Schmiedt, R. A. (1992). Lateral wall Na,K-ATPase and endocochlear potentials decline with age in quiet-reared gerbils. Hear Res, 61(1-2), 35–46. 10.1016/0378-5955(92)90034-k

50. Seabold, S., & Perktold, J. (2010). Statsmodels: Econometric and statistical modeling with python. Proceedings of the 9^th^ Python in Science Conference. 10.25080/Majora-92bf1922-011

51. Sergeyenko, Y., Lall, K., Liberman, M. C., & Kujawa, S. G. (2013). Age-related cochlear synaptopathy: an early-onset contributor to auditory functional decline. J Neurosci, 33(34), 13686–13694. 10.1523/JNEUROSCI.1783-13.2013

52. Shaheen, L. A., Valero, M. D., & Liberman, M. C. (2015). Towards a Diagnosis of Cochlear Neuropathy with Envelope Following Responses. J Assoc Res Otolaryngol, 16(6), 727–745. 10.1007/s10162-015-0539-3

53. Suthakar, K., & Liberman, M. C. (2021). Auditory-nerve responses in mice with noise-induced cochlear synaptopathy. J Neurophysiol, 126(6), 2027–2038. 10.1152/jn.00342.2021

54. Takahashi, T. (1971). The ultrastructure of the pathologic stria vascularis and spiral prominence in man. Ann Otol Rhinol Laryngol, 80(5), 721–735. 10.1177/000348947108000515

55. Trune, D. R., Mitchell, C., & Phillips, D. S. (1988). The relative importance of head size, gender and age on the auditory brainstem response. Hear Res, 32(2), 165–174.

56. Vaden, K. I., Jr., Eckert, M. A., Matthews, L. J., Schmiedt, R. A., & Dubno, J. R. (2022). Metabolic and Sensory Components of Age-Related Hearing Loss. J Assoc Res Otolaryngol, 23(2), 253–272. 10.1007/s10162-021-00826-y

57. Van Der Biest, H., Keshishzadeh, S., Keppler, H., Dhooge, I., & Verhulst, S. (2023). Envelope following responses for hearing diagnosis: Robustness and methodological considerations. J Acoust Soc Am, 153(1), 191. 10.1121/10.0016807

58. Vasilkov, V., Garrett, M., Mauermann, M., & Verhulst, S. (2021). Enhancing the sensitivity of the envelope-following response for cochlear synaptopathy screening in humans: The role of stimulus envelope. Hear Res, 400, 108132. 10.1016/j.heares.2020.108132

59. Verhulst, S., Altoe, A., & Vasilkov, V. (2018). Computational modeling of the human auditory periphery: Auditory-nerve responses, evoked potentials and hearing loss. Hear Res, 360, 55–75. 10.1016/j.heares.2017.12.018

60. Verhulst, S., Dau, T., & Shera, C. A. (2012). Nonlinear time-domain cochlear model for transient stimulation and human otoacoustic emission. J Acoust Soc Am, 132(6), 3842–3848. 10.1121/1.4763989

61. Verhulst, S., Jagadeesh, A., Mauermann, M., & Ernst, F. (2016). Individual Differences in Auditory Brainstem Response Wave Characteristics: Relations to Different Aspects of Peripheral Hearing Loss. Trends Hear, 20. 10.1177/2331216516672186

62. Wang, Y., Fallah, E., & Olson, E. S. (2019). Adaptation of Cochlear Amplification to Low Endocochlear Potential. Biophys J, 116(9), 1769–1786. 10.1016/j.bpj.2019.03.020

63. Wilson, J. L., Abrams, K. S., & Henry, K. S. (2021). Effects of Kainic Acid-Induced Auditory Nerve Damage on Envelope-Following Responses in the Budgerigar (Melopsittacus undulatus). J Assoc Res Otolaryngol, 22(1), 33–49. 10.1007/s10162-020-00776-x

64. Wu, P. Z., Liberman, L. D., Bennett, K., de Gruttola, V., O’Malley, J. T., & Liberman, M. C. (2019). Primary Neural Degeneration in the Human Cochlea: Evidence for Hidden Hearing Loss in the Aging Ear. Neuroscience, 407, 8–20. 10.1016/j.neuroscience.2018.07.053

65. Wu, P. Z., O’Malley, J. T., de Gruttola, V., & Liberman, M. C. (2020). Age-Related Hearing Loss Is Dominated by Damage to Inner Ear Sensory Cells, Not the Cellular Battery That Powers Them. J Neurosci, 40(33), 6357–6366. 10.1523/JNEUROSCI.0937-20.2020

66. Wu, P. Z., O’Malley, J. T., de Gruttola, V., & Liberman, M. C. (2021). Primary Neural Degeneration in Noise-Exposed Human Cochleas: Correlations with Outer Hair Cell Loss and Word-Discrimination Scores. J Neurosci, 41(20), 4439–4447. 10.1523/JNEUROSCI.3238-20.2021

67. Zhu, L., Bharadwaj, H., Xia, J., & Shinn-Cunningham, B. (2013). A comparison of spectral magnitude and phase-locking value analyses of the frequency-following response to complex tones. J Acoust Soc Am, 134(1), 384–395. 10.1121/1.4807498

